# Statistical uncertainty explains the poor agreement in polygenic scoring for type 2 diabetes

**DOI:** 10.64898/2026.02.25.26347015

**Authors:** Ravi Mandla, Xinzhe Li, Zhuozheng Shi, Sarah A. Abramowitz, Sandra Lapinska, Penn Medicine Biobank, Michael G. Levin, Scott M. Damrauer, Bogdan Pasaniuc

## Abstract

Polygenic scores (PGS) have emerged as an important tool for genetic risk prediction in medicine to identify individuals at high-risk for disease. A major limitation in their implementation is the apparent disagreement among scores for the same individual decreasing their interpretability and utility in clinical settings. Here we show that the poor agreement across PGSes for type 2 diabetes (T2D) is fully explained by statistical uncertainty in PGS-based prediction; individual-level uncertainty estimates from a single PGS explain the variability across existing PGSes. We provide an approach for the selection of high-risk individuals that incorporates measures of uncertainty and show that individuals with high confidence based on their PGS uncertainty have higher risk agreement across existing PGS and are more likely to develop T2D than high-risk individuals based on only point estimates of PGS. Together, these findings shed light on the factors underlying a roadblock in PGS implementation and underscore the need to incorporate uncertainty in PGS-based predictions.

## Introduction

Diabetes places an enormous burden on healthcare systems, accounting for 20% of in-patient visits within the United States^1^. Furthermore, this burden is likely to increase, with an estimated 246 million additional individuals globally developing diabetes in the next 20 years^2^. The early detection of type 2 diabetes (T2D) may significantly reduce this burden through early intervention strategies to prevent T2D and related complications^2,3^. To this end, polygenic scores (PGS) have emerged as a potential viable tool to identify individuals at risk of developing T2D based on their genetics^4,5^. Yet, the adoption of T2D PGS into routine care is limited due to their overall lower accuracy compared to clinical risk factors^6^, their unclear impacts on improving disease treatment^7^, and their poor portability between populations^8,9^. Nevertheless, T2D PGS are strongly associated with disease incidence^10,11^ and have been shown to provide additional disease predictive information over traditional clinical risk factors^11,12^, highlighting their potential in improving T2D onset prediction.

A critical limitation in implementing genetic prediction for T2D is that different PGS often disagree with each other at an individual-level on a patient’s risk for disease^13–16^. For example, 20% of individuals placed in the top 5% of risk based on one coronary heart disease (CHD) PGS were also placed in the bottom 5% of risk based on a separate CHD PGS^15^. While strategies have been proposed to mitigate this disagreement in risk^16^, it still remains unclear why this disagreement exists and varies substantially between individuals. As such, it is difficult to determine the best practices to address this individual-level disagreement and thus implement PGS in clinical settings to identify patients with truly high genetic risk^17^.

In this work, we characterize the concordance across published T2D PGS and examine potential factors that may underly the poor agreement between different PGS for T2D. We identify statistical uncertainty in PGS estimation, from the selection and weighting of genetic variants, as the main factor contributing to lack of agreement in T2D PGS risk estimation. We demonstrate that the statistical uncertainty, as estimated using Bayesian-based approaches, can be leveraged to improve the identification of individuals with high PGS for T2D. Consequently, these individuals also have the highest risk agreement across PGSes, so their PGS risk estimates may be most clinically actionable.

## Results

### Overview of individual-level PGS uncertainty

Various existing approaches for PGS construction, including LDPred2 and PRS-CS, provide standard errors for PGS weights that can be used to estimate confidence intervals for PGSs at an individual level^13,18,19^. Consequently, individuals above a PGS threshold *t* based only on their PGS point estimate may be further stratified based on their individual-level probability of having their entire PGS credible interval above *t*. We refer to this individual-level probability as “PGS high risk confidence” (**Figure 1A**). As an illustrative example, consider 10 published PGSes for a given condition with similar population-level accuracy, each applied to three individuals of varying genetic risk (**Figure 1B**). Each participant may have both different averages and variances across all ten PGS based on their unique genotype values and PGS weights. If statistical uncertainty in PGS weight estimation is main factor in variability of prediction across PGSes, we expect the prediction interval estimated by a single PGS to approximate variability across all PGSes (**Figure 1BC**; **Methods**). Deviations from the y=x line suggest poor calibration of the PGS individual measures of uncertainty in explaining variability across PGSes, suggesting that other factors contribute to PGS disagreement (**Supplementary Figure 1**; **Methods**).

**Figure 1:**
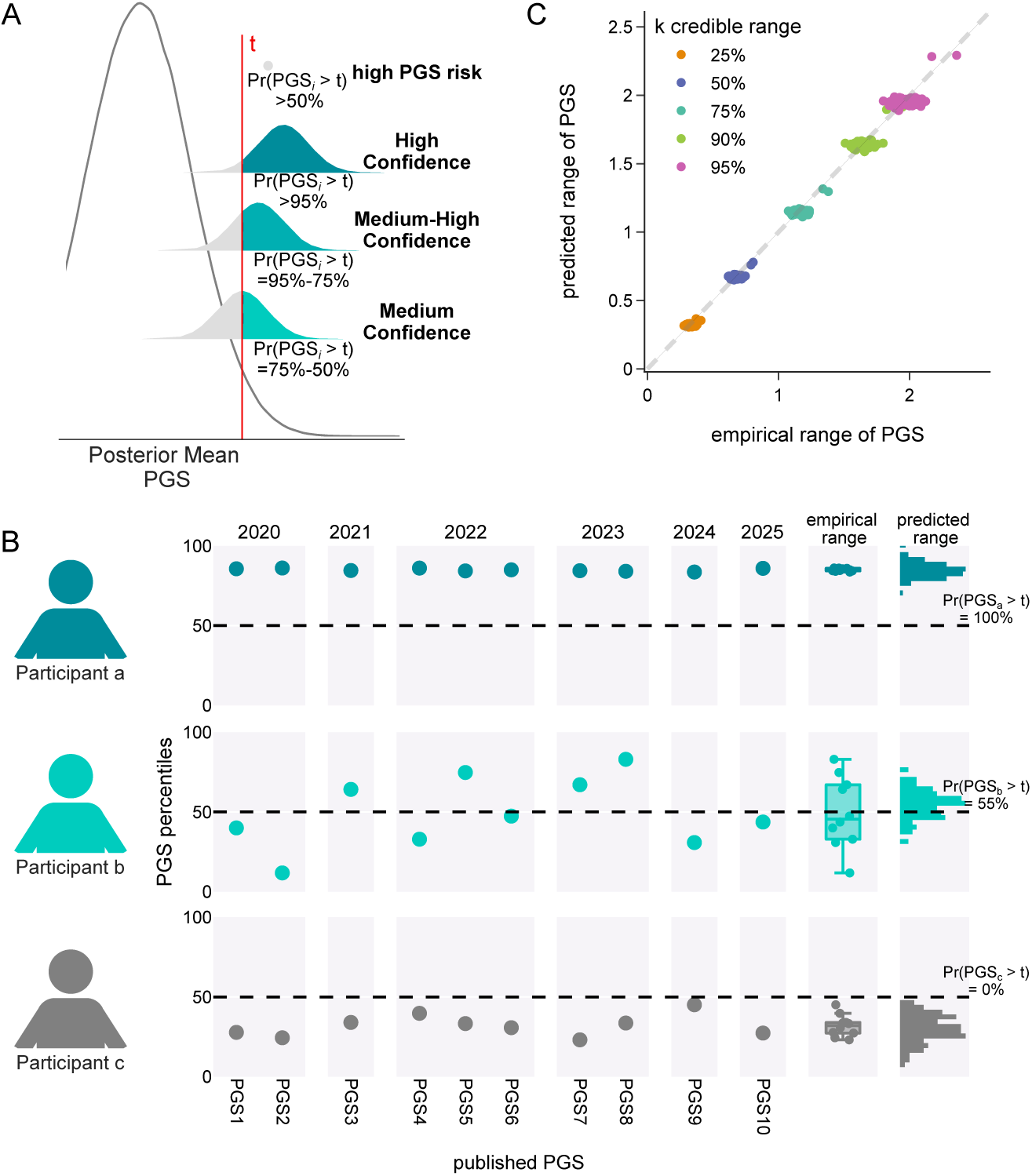
Overview of the relationship between predicted uncertainty in PGS and empirical variance across published PGS. (**A**) Individuals are traditionally called as high risk if their PGS is above a threshold t. With predicted uncertainty information, individuals can be further stratified based on their confidence for being at high risk, defined as the proportion of their posterior PGS distribution over the risk threshold. (**B**) In a toy example of three participants with differing averages and variance across 10 published PGS with similar population-level accuracy, the empirical range estimated from these 10 PGS correlates with the predicted range based on the uncertainty in a single, independent PGS. (**C**) The predicted range based on a single PGS will approximate the empirical range of multiple PGS for a variety of range sizes (25%, 50%, 75%, 90%, 95% credible ranges).

### T2D PGS uncertainty approximates disagreement across published T2D PGSes

To evaluate the agreement across PGS for T2D, we first extracted electronic health record (EHR) and genetic data from 223,015 participants within the v7 data release of the *All of Us Research Program* (AoU), including 24,395 individuals with T2D and 198,080 individuals without evidence of diabetes (**Methods**, **Supplementary Table 1**). As AoU participants reflect the diverse communities of the US, and since PGS are known to perform differently across different populations^9,20,21^, we initially restricted our analysis to 118,058 individuals genetically similar to European ancestries (**Methods**). We refer to these individuals as EUR-like, following previous guidelines on classifying individuals based on genetic similarity^22,23^. We next calculated 57 different T2D PGS per individual using published weights from the PGScatalog (**Methods**, **Supplementary Table 2**). As comparator, we estimated a new T2D PGS using PRS-CS^19^ and the current largest T2D GWAS meta-analysis of EUR-like individuals^24^ (T2DGGI-EUR; **Methods**). For conservativeness, we restricted the published PGS to the five scores with similar population-level accuracy based on a region of practical equivalence (ROPE) +/- 0.02 around the area under the receiver operator curve (AUC)^15^ to the T2DGGI-EUR score (**Supplementary Figure 2**; **Supplementary Table 3**; **Methods**).

We observed a large disagreement in risk across the six T2D PGS, with an average pairwise risk concordance of 0.82, ranging from 0.73 to 0.90 (**Supplementary Figure 3**; **Methods**). 6,379 (5.4%) of individuals were above the risk cutoff of top 2%^25^, as recommended by the eMERGE consortium, in at least one of these six PGS. However, 2,915 (45.7%) of these individuals were only in the top 2% of one of these PGS, highlighting overall poor risk agreement at individual level between these different T2D PGS (**Supplementary Figure 4**). Sixty-seven (1.1%) of these individuals were in the top 2% of one PGS but in the bottom 50% of another PGS, while no individuals were in the top 2% of one PGS but in the bottom 2% of another PGS. In sensitivity analyses, we found increasing the ROPE stringency to +/- 0.01 identified only one published PGS with population-level equivalence to the T2DGGI-EUR score, while decreasing the stringency to +/- 0.04 identified three additional PGS (**Supplementary Figure 5A, B**; **Supplementary Table 3**). The average pairwise risk concordance also decreased to 0.78 when using a ROPE +/- 0.04 (**Supplementary Figure 5C**). We also tested how the number of individuals classified as high risk across multiple PGS changes with different high-risk cutoffs. We found that using a more lenient risk cutoff of top 10% increased risk agreement, with 24,919 individuals identified as high risk based on any PGS while 8,075 (32.4%) of them were high risk based on only one PGS (**Supplementary Figure 6**).

Having confirmed the lack of agreement among PGSes for T2D, we next asked whether individual-level uncertainty in PGS (as estimated using PRS-CS for T2DGGI-EUR PGS) can explain the disagreement among all other PGSes for T2D. We found that the empirical prediction range (defined as the empirical standardized range for a given individual across the five published PGSes, **Methods**) is equivalent with the prediction range as estimated by the T2DGGI-EUR PGS. This shows that disagreement among PGS individual-level predictions for T2D are due to statistical noise in the estimation of PGS weights and this disagreement can be estimated based only on a single PGS (slope = 0.96; **Figure 2A**). We replicated these results using EUR-like individuals in the Penn Medicine Biobank (**Supplementary Table 4**), finding similar results (slope = 0.97; **Supplementary Figure 7**; **Methods**). To determine that our results are not due to choice of PGSes for T2D, we recalculated the empirical range sizes when using alternative ROPE cutoffs. As expected, decreasing stringency on defining population-level equivalence PGS to a ROPE +/- 0.04 also decreased the calibration between predicted and empirical range lengths (slope = 0.89) due to the increased empirical range length (**Supplementary Figure 8**). We could not test the change in relationship between the predicted and empirical ranges with a more stringent ROPE +/- 0.01 due to only one published PGS surviving this strict filtering step (**Supplementary Figure 5A**; **Supplementary Table 3**).

**Figure 2:**
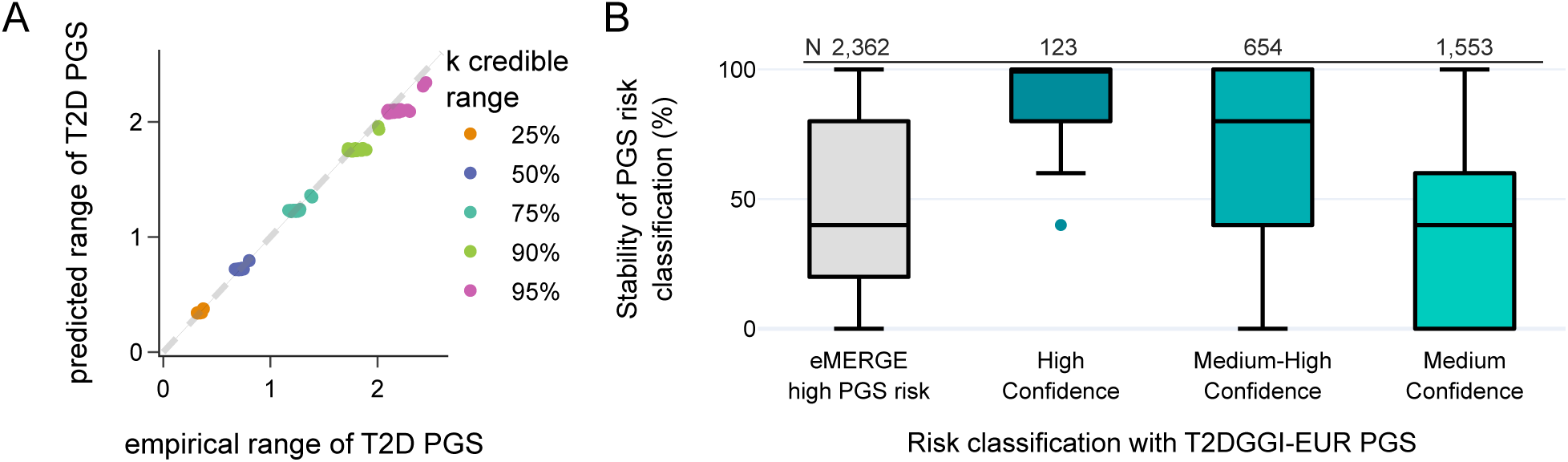
PGS risk confidence information stratifies individuals with differing risk agreement. (**A**) Scatterplot of the empirical range of T2D PGS based on published scores to predicted range based on the T2DGGI-EUR score from PRS-CS, across different credible range sizes. Due to the small number of published T2D PGS used, we grouped individuals into percentile bins prior to range calculation (**Methods**). PGS are all standardized prior to range estimation. (**B**) Comparison of risk stability across individuals identified using the eMERGE risk cutoff of top 2% with the T2DGGI-EUR score and individuals identified at different confidence classifications with the T2DGGI-EUR score. Stability per individual was calculated as the average number of PGS that call said individual as high risk. High confidence refers to >95% probability of PGS being over the risk threshold, Medium-high confidence refers to >75% and <=90% probability of PGS being over the risk threshold, and Medium confidence refers to >50% and <=75% probability of PGS being over the risk threshold.

### Incorporation of PGS uncertainty improves risk classification for T2D

Having showed that confidence intervals of the T2DGGI-EUR PGS approximated the empirical range across other five published T2D PGSes, we next hypothesized that leveraging uncertainty in PGS-based prediction can identify individuals with the highest risk agreement (**Figure 1A**). Focusing on the 2,137 individuals at top 2% of T2DGGI-EUR PGS, 123 had a >95% confidence of being above the risk threshold. The vast majority of them showed high degrees of agreement, with on average 89% of the other five PGS also calling them high risk (**Figure 2B**). Similar results were observed at lower confidence levels, with a tight coupling between confidence levels and agreement across scores e.g., medium-high (N=654; 66.4%; Mann Whitney U P = 9.3×10^-16^) and medium confidence groups (N=1,553; 38.5%; Mann Whitney U P = 3.1×10^-49^; **Figure 2B**; **Supplementary Table 5**). Similar results were obtained at high-risk cutoff of 10% (**Supplementary Figure 9**; **Supplementary Table 6**). Recent publications have introduced novel approaches for estimating PGS prediction intervals^21,26,27^. We evaluated PredInterval^27^ on our data to find that PredInterval estimated ranges two times larger than PRS-CS leading to mis-estimation of the variability across other PGS scores (**Supplementary Figure 10**). Most notably, PredInterval did not identify any individual having high confidence for being in the high-risk category due to its large prediction intervals.

Next we evaluated the impact of choice of PGS on our results using a new multi-ancestry PGS for T2D from five single population GWAS meta-analyses^24,28^ (T2DGGI-MA PGS; **Methods**). Overall, the T2DGGI-MA PGS had a smaller 95% credible range compared to the T2DGGI-EUR PGS, likely owing to its larger sample size (**Supplementary Figure 11A**). Importantly, we found wide overlap between the credible ranges of the T2DGGI-MA and T2DGGI-EUR scores for most people. On average, 90% of the T2DGGI-MA posterior samples were within the 95% credible range of the T2DGGI-EUR score across all EUR-like individuals and only 725 (0.6%) individuals had less than 50% of their T2DGGI-MA posterior samples within their T2DGGI-EUR 95% credible range (**Supplementary Figure 11B**). Furthermore, we found that 67 (54%) of individuals identified as high confidence in the T2DGGI-EUR score also had high risk confidence in the T2DGGI-MA score. However, 114 (93%) had at least medium-high risk confidence in the T2DGGI-MA score (**Supplementary Figure 11C**), demonstrating that classifications of risk confidence labels may change between PGS.

Individuals with high confidence of being at the top percentiles may also have a higher PGS point estimates compared to individuals with medium-high or medium confidence. Indeed, we found strong positive correlation between the point estimates and risk confidence values of the T2DGGI-EUR score (Spearman R = 0.98, P < 10^-300^; **Supplementary Figure 12A**). We next compared whether individuals identified as high confidence will always be identified as having the highest PGS point estimate. When taking the top 123 individuals with the highest PGS, we found large overlaps with the 123 high confidence individuals (**Supplementary Figure 12B**). Individuals identified only from this rank-order based approach had similar risk agreement to individuals identified only as high confidence (Mann-Whitney U P = 0.81; **Supplementary Figure 12C**). This suggests that individuals with the highest agreement of being high risk tend to also have the highest PGS.

### Risk confidence estimation generalizes across population groups

Previous work has demonstrated that PGS population-level and individual-level accuracy varies between population groups^9,29^, though it is unknown whether individual-level risk agreement varies as well. We extended our analyses across all 223,015 participants of various genetic ancestries in All of Us, focusing on the T2DGGI-MA PGS. After filtering PGS to those with similar population-level accuracy in every genetically inferred population group, we were left with seven total published PGS, including the T2DGGI-MA PGS and six other published scores (**Supplementary Figure 13**; **Supplementary Table 7**; **Methods**). Individual-level agreement across these seven PGS was worse in individuals genetically similar to African populations (AFR-like), with an average rank concordance of 0.67, compared to EUR-like individuals, with an average rank concordance of 0.80 (**Supplementary Figure 14**). Furthermore, 1,193 (63.4%) of the 1,881 AFR-like individuals above the top 2% risk threshold in one PGS were below the same threshold in all other PGS, compared to 3,435 (43.6%) of the 7,878 EUR-like individuals.

We next evaluated whether the T2DGGI-MA PGS posterior distribution approximated the empirical distribution across published PGS in the multi-ancestry setting. We found overall consistency in the ranges between the predicted range from the T2DGGI-MA score and the empirical range across populations (**Figure 3A**). However, the empirical range was consistently larger than the predicted range (slope in all participants = 0.81), suggesting potential under-calibration of the T2DGGI-MA PGS posterior distribution for approximating the published PGS. We then identified all individuals called as high risk based on the T2DGGI-MA PGS with the eMERGE risk cutoff of top 2%. Risk stability across the published PGS was overall low, with a participant in the top 2% of the T2DGGI-MA PGS only being in the top 2% of 40.0% of the six published PGS on average. This stability was highest for EUR-like individuals (49.1%) and lowest for AFR-like individuals (17.7%) (**Figure 3B**).

**Figure 3:**
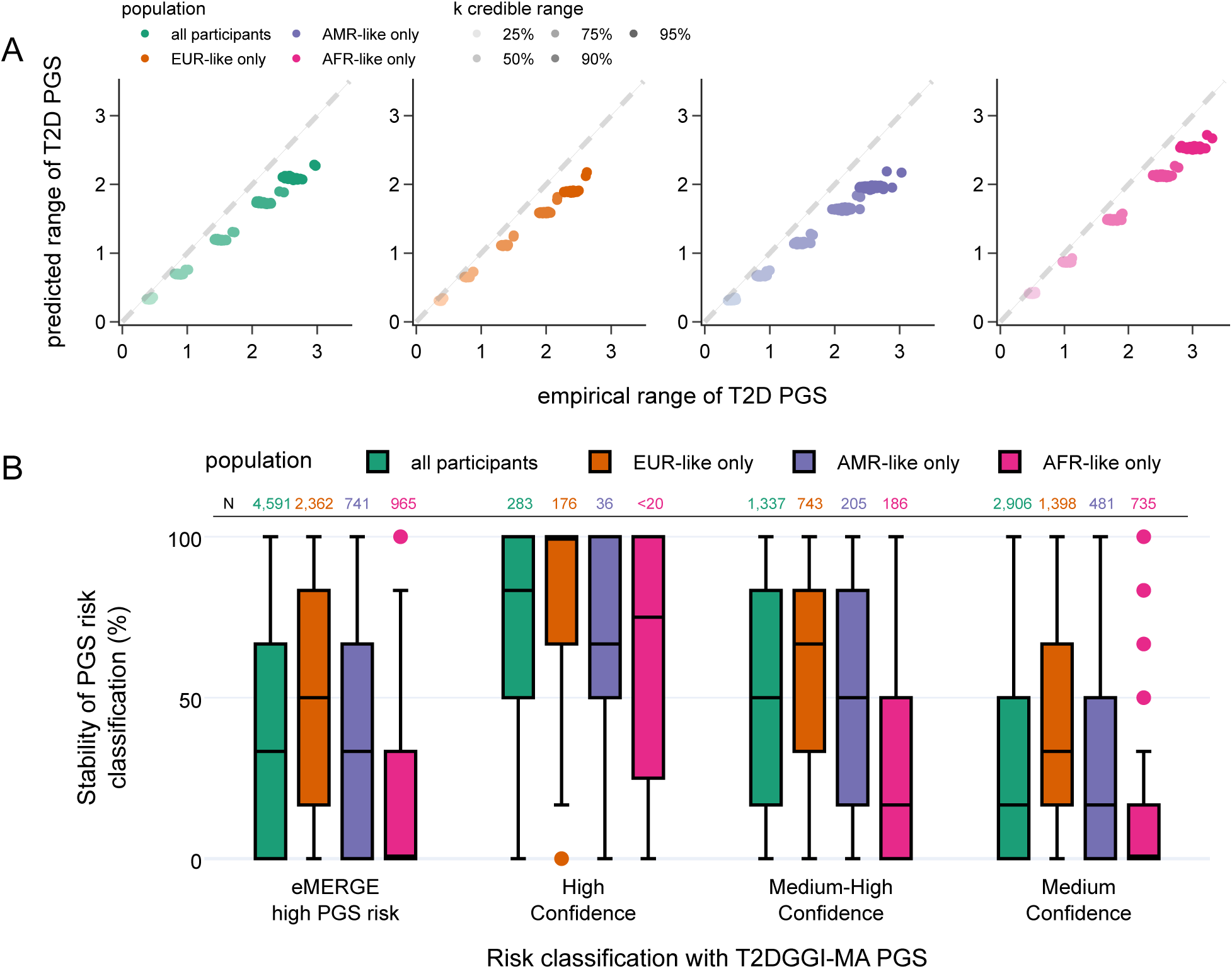
PGS risk agreement and confidence varies by population. (**A**) Scatterplot of the empirical range of T2D PGS based on published scores to predicted range based on the T2DGGI-MA score from PRS-CSx, across different credible range sizes and different population groupings (**Methods**). Colors indicate population group stratifications using genetic similarity, while intensity of color indicates credible range grouping. (**B**) Comparison of risk stability across individuals identified using the eMERGE risk cutoff of 0.02 based on the T2DGGI-MA score and individuals identified at different confidence classifications based on the T2DGGI-MA score. Stability per individual was calculated as the average number of PGS that call said individual as high risk. Colors indicate population group stratifications using genetic similarity. EUR-like only: genetically similar to European populations. AMR-like only: genetically similar to Admixed American populations. AFR-like only: genetically similar to African populations.

We further classified these individuals into risk confidence bins as done previously with the T2DGGI-EUR score. In total, 283 of EUR-like individuals were identified as high confidence, with an average risk stability of 81.3%. In contrast, a much smaller number of AFR-like individuals (<20) were identified as high confidence, with an average risk stability of 65.3%. Overall, individuals with high confidence of being high risk had significantly higher risk stability across each population groups relative to individuals with medium-high or medium confidence, with this stability highest for EUR-like individuals and lowest for AFR-like individuals (**Figure 3**; **Supplementary Table 8**). These results underscore a potential for increasing health inequity when incorporating measures of uncertainty in prediction of high-risk individuals using PGS.

### Risk confidence associates with T2D onset

We hypothesized that individuals with high confidence for being high risk may also be more likely develop T2D. To this end, we subset AoU to all individuals with a recorded outpatient visit with a clinician and compared time of T2D onset between individuals of varying risk confidence groupings (N=142,536; **Supplementary Table 9**; **Methods**). To increase power, we also re-defined risk confidence using a PGS risk cutoff of top 10%. Indeed, these high confidence individuals within AoU had higher T2D incidence rates (33.1%) compared to medium-high (26.5%; χ^2^ P = 2.3×10^-6^; log-rank P = 1.3×10^-6^) and medium (23.7%; χ^2^ P = 1.6×10^-13^; log-rank P = 2.5×10^11^) confidence groups (**Figure 4A**; **Supplementary Table 9**). High confidence individuals were also enriched for EUR-like ancestry, had a lower age to T2D onset, and had both higher family history of T2D and random glucose compared to medium-high and medium confidence individuals (**Supplementary Table 9**; **Methods**). Furthermore, high confidence individuals had a higher absolute probability of developing T2D, based on both genetics and relevant clinical risk factors^30^, compared to individuals with medium confidence (23.0% vs 20.8%; Mann-Whitney U P = 0.03; **Figure 4B**; **Methods**). To test if information on PGS risk confidence could improve the identification of individuals likely to develop T2D in clinical settings, we next created survival models consisting of relevant clinical risk and the T2DGGI-MA PGS, with or without risk confidence information (**Methods**). As risk confidence and PGS are highly correlated (**Supplementary Figure 12**), risk confidence was unsurprisingly non-significant after adjusting for the T2DGGI-MA PGS and other relevant clinical risk factors (**Supplementary Figure 15**; **Supplementary Table 10**). Overall, these findings highlight how stratifications of participants into confidence groupings may identify individuals with higher onset rates, though this information may not improve disease prediction over existing genetic models.

**Figure 4:**
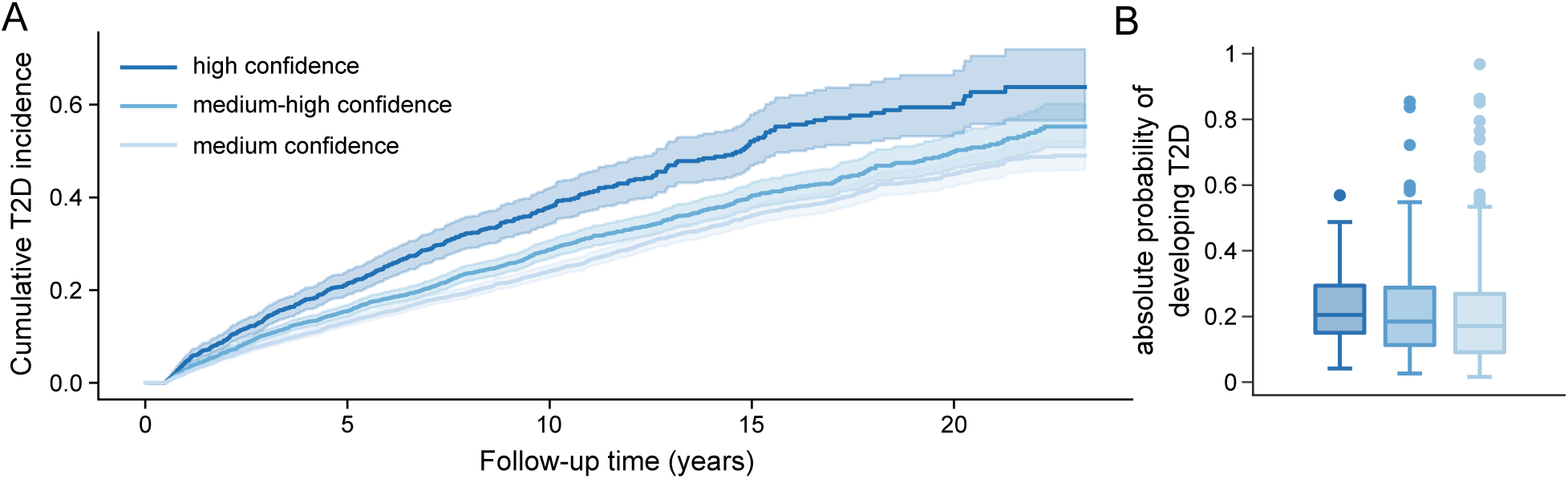
PGS risk confidence predicts onset of T2D. (**A**) Cumulative incidence curves of T2D for individuals identified as high, medium-high, or medium confidence for being over the risk threshold (top 10%). (**B**) Absolute risk of developing T2D over the follow-up time, stratified by confidence. Absolute risk was calculated using a logistic regression model including clinically risk factors^30^, the T2DGGI-MA PGS, and risk confidence information as a quantitative value. Clinical risk factors included age, sex, BMI, smoking status, family history of T2D, systolic blood pressure, random glucose, total cholesterol, high-density lipoprotein cholesterol, and triglycerides.

### Statistical uncertainty explaining PGS agreement is trait specific

To test the generalizability of our approach to other traits, we repeated the above analyses for two additional traits: body mass index (BMI) and CHD (**Methods**). We estimated new PGS using PRS-CS and high powered GWAS summary statistics for BMI^31^ and CHD^15^, calculated PGS for BMI and CHD using published weights from PGScatalog, and filtered the published scores to those with ROPE +/- 0.02 to the AUC of the newly estimated PGS (**Methods**). In total, this left us with 19 published BMI PGS and 39 published CHD PGS (**Methods**; **Supplementary Figure 16, 17; Supplementary Table 11-14**). For consistency, we also performed this analysis only in EUR-like individuals. Like T2D, the predicted range of the BMI PGS approximated the empirical range across published scores (slope = 1.0; **Figure 5A**, left panel). Individuals in the eMERGE recommended risk cutoff of top 3%^25^ based on our BMI PGS, on average, were also called as high risk in 53.2% of the other 19 PGS. After stratifying individuals into confidence groupings, individuals with high confidence were called as high risk in 92.4% of the other published BMI PGS, significantly higher than individuals with medium-high (71.1%; Mann-Whitney U P = 4.1×10^-44^) and medium confidence (42.9%; Mann-Whitney U P = 1.1×10^-101^; **Figure 5B**, right panel; **Supplementary Table 15**).

**Figure 5:**
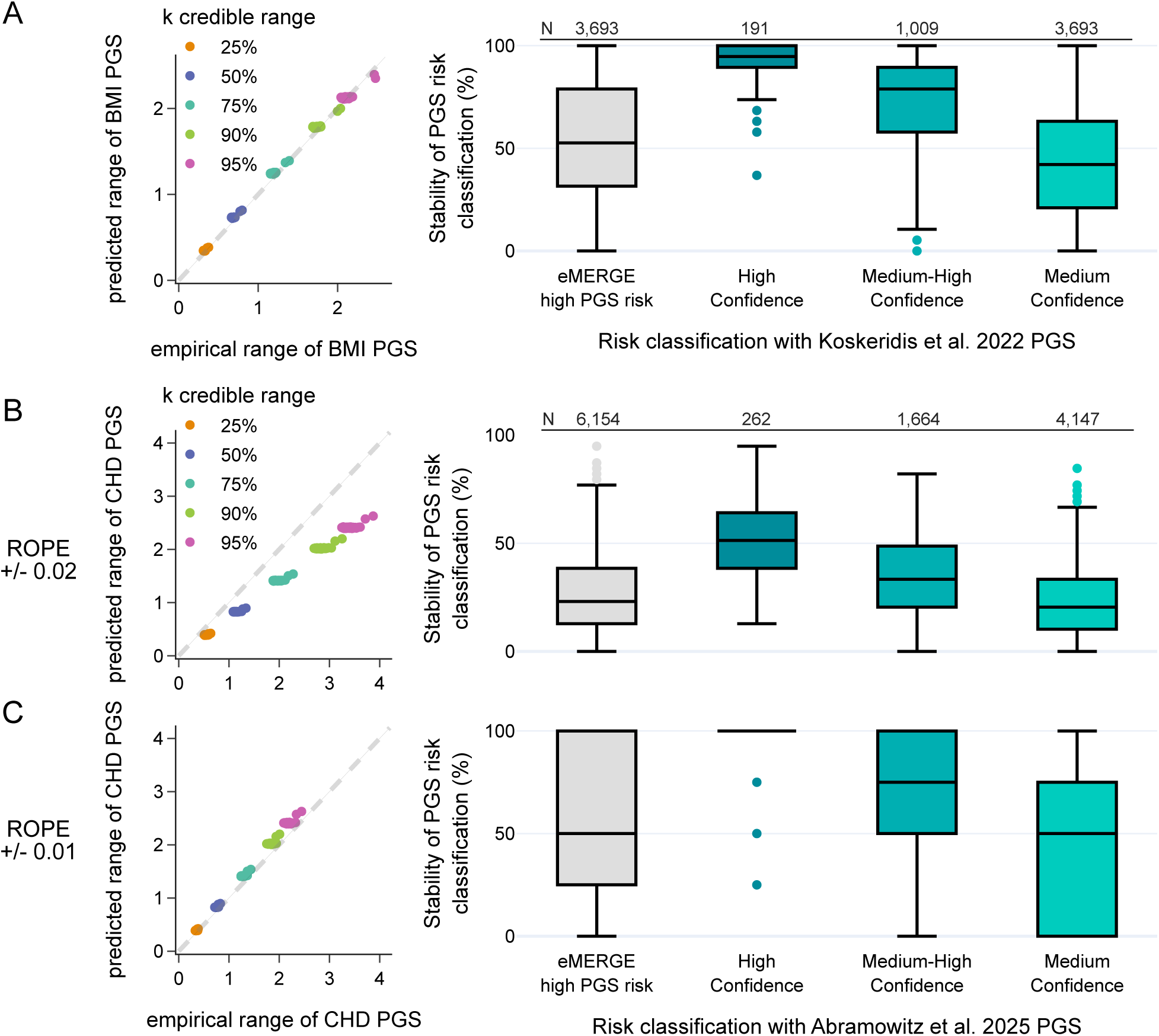
PGS uncertainty in BMI and CHD approximates differing PGS risk agreements for both traits. (**A**) **Left panel:** Scatterplot of the empirical range of body mass index (BMI) PGS based on published scores to predicted range based on a novel score calculated with PRS-CS (Koskeridis et al. 2022), across different credible range sizes (**Methods**). **Right panel:** Comparison of risk stability across individuals identified using the eMERGE risk cutoff of 0.03 with the Koskeridis et al. 2022 score and individuals identified at different confidence classifications with the Koskeridis et al. 2022 score. (**B**)-(**C**) depict similar results for coronary heart disease (CHD) when using either published, population equivalent PGS defined using a (**B**) ROPE +/- 0.02 or (**C**) +/- 0.01. Both (**B**) and (**C**) estimate the predicted range based on a novel score calculated with PRS-CS (Abramowitz et al. 2025). Stability per individual was calculated as the average number of PGS that call said individual as high risk.

Interestingly, we found miscalibration of the predicted range of the CHD PGS, with overall larger empirical ranges and lower risk stability compared to T2D and BMI (slope = 0.71; **Figure 5B**, left panel). For example, high confidence individuals based on our PGS were only above the eMERGE recommended risk cutoff of top 5%^25^ in 50.9% of the other 39 CHD PGS, though this was still significantly higher than medium-high (35.1%; Mann-Whitney U P = 3.3×10^-36^) or medium confidence (22.9%; Mann-Whitney U P = 9.3×10^-97^) individuals (**Supplementary Table 16**). However, further restricting the published PGS using a stringent ROPE of +/- 0.01, for a total of four published CHD PGS, improved the calibration of the predicted range by decreasing the empirical range (slope = 1.1; **Figure 5C**, left panel). As such, stability overall increased for every individual, with high confidence individuals now on average above the recommended risk cutoff in 95.2% of these four published CHD PGS (**Figure 5C**, right panel; **Supplementary Table 16**). These findings highlight the overall calibration of uncertainty of a single PGS to approximate varying risk across published PGS for different traits, with calibration worse for certain diseases.

## Discussion

We showed that the disagreement in PGS-based risk estimates for T2D is due to statistical noise and we demonstrated that individual statistical uncertainty of a single PGS for T2D is sufficient to explain the variability across all PGS scores for T2D. We also showed that approaches which take PGS uncertainty into account when stratifying individuals as high risk identify individuals more likely to develop T2D than approaches based on point estimates of T2D PGS alone. However, as risk confidence information was highly correlated with the PGS point estimates themselves, there may be minimal improvements in overall disease prediction at the population level. Nevertheless, this risk confidence information is useful for informing which individuals have high confidence to be at tails of the genetic risk distribution using PGS. Alternative methods for conveying PGS risk uncertainty, for example returning credible ranges of the PGS risk, may also help in identifying patients who have the lowest uncertainty in their PGS. Future work is required to investigate how clinicians and patients interpret these estimates, and how these estimates may inform clinical care. Furthermore, since individual-level PGS uncertainty is correlated with genetic distance to the majority population included in the GWAS^29^, AFR-like individuals have larger uncertainty in their PGS and thus are underrepresented in high risk confidence groups. Thus, further work is also required to assess the potential health inequities that PGS uncertainty information may introduce into clinical care.

Beyond T2D, we found that uncertainty in a single PGS approximated poor risk agreement in published PGS for other cardiometabolic traits as well, including BMI and CHD. This suggests that inherent differences in variant weighting underlies poor risk agreement of PGS at an individual-level across many traits. However, calibration was overall lower for CHD than BMI and T2D, with proper calibration for CHD only reached after stringent filtering of scores for population-level equivalence. This worse performance for CHD may be explained by the greater amount of missing heritability in CHD GWAS compared to T2D GWAS, with up to 26% missing heritability in CHD^32^ compared to 10% in T2D^33^.

Accounting for the poor individual-level agreement of risk is a major challenge in deploying PGS into clinical settings^17^, particularly because it remained unclear why this poor agreement exists. The only current approach to minimize this poor agreement is to perform a weighted sum across PGS, iteratively adding them into the “final” score in order of publication date^16^. However, it is unclear how these weights will change as the training data used for estimating the weights changes. Furthermore, this approach will inherently ignore potential differences in PGS variance between individuals, leading to similar risk classification for individuals with the same estimated PGS but with different variance around the estimate. Here, we found that individual-level uncertainty inherent in PGS estimation explains the variance in published PGS, thus providing a solution towards incorporation of uncertainty using a single (most accurate) PGS together with its estimates of uncertainty.

In this work, we estimated individual-level uncertainty from a PGS under the assumptions of the PRS-CS model, however updated methods for estimating PGS uncertainty may improve the capture of individuals with true high genetic risk for disease^26,27^. We evaluated one of these approaches, PredInterval, which estimates the individual-level prediction interval of a PGS based on its overall population-level performance. Prediction intervals from PredInterval were much larger than those estimated from PRS-CS, suggesting that PredInterval is mis-calibrated for approximating published PGS risk agreement. QR-PRS^26^, another approach which estimates uncertainty in PGS based on quantile regression, may also improve calibration over the PRS-CS model, albeit QR-PRS is only designed currently for non-disease continuous traits.

Our work has several limitations. First, we tested the relationship between predicted PGS credible ranges and empirical PGS ranges using the cardiometabolic traits T2D, BMI, and CHD. However, the relationship between these ranges may differ as the genetic architecture of the analyzed traits vary. Consequently, future work inspecting this relationship for a broader, more diverse number of traits is necessary to better understand the generalizability of individual-level PGS uncertainty to approximate observed poor agreement in PGS risk. Second, we estimated individual-level uncertainty in PGS using PRS-CS and PRS-CSx, though this estimated uncertainty may change between different methods. Third, we note that our metrics of calibration are dependent on how we defined population-level equivalence across published PGS, with notable differences in performance for CHD based on the ROPE range. However, for T2D we found that calibration of the predicted and empirical ranges were more stable, with similar overall calibration when using a more lenient ROPE range of 0.04. Finally, we tested for the association of PGS risk confidence and onset of T2D among individuals with recorded primary care visits in AoU. We acknowledge, however, that missing data and inaccuracies in ICD-based identification of T2D status may lead to inaccurate time-to-onset measurements and reduced model accuracy.

In summary, we found that individual-level uncertainty in a single T2D PGS approximated variance across published PGS, highlighting the role that uncertainty in variant weighting plays in observed PGS risk stability. Given this, reporting uncertainty in PGS may improve their utility both through the identification of individuals with robust risk estimates and the selection of PGS with both high population-level and individual-level accuracy.

## Methods

### Study Population

We performed our primary analyses using data from the *All of Us Research Program* (AoU), a large, hospital system-based Biobank containing short read whole genome sequencing (srWGS) data, survey information, and electronic health record (EHR) data from adults across the United States. We pulled data from AoU controlled tier v7 for our primary analyses.

We performed replication analyses using data from the *Penn Medicine Biobank*, which collects genetic data and electronic health record data from consenting individuals within the University of Pennsylvania Health System.

### Phenotype Data Curation

We extracted lab measurements for random glucose using LOINC codes 2345-7 in units mg/dL, systolic blood pressure using LOINC codes 8480-6 in mmHg, total cholesterol using LOINC codes 2093-3 in mg/dL, high-density lipoprotein (HDL) cholesterol using LOINC codes 2085-9 in mg/dL, and triglycerides using LOINC codes 2571-8 in mg/dL. We also extracted BMI values in units kg/m2. For all quantitative traits, we removed outlier values, defined as measurements being over four standard deviations from the mean in log scale. We then defined type 2 diabetes (T2D) prevalence cases and controls using a previously established EHR-based phenotyping algorithm leveraging these cleaned lab values in-tandem with ICD-codes for diabetes and medication records^34^ and we defined T2D incidence cases using only the ICD-codes. We defined coronary heart disease (CHD) cases based on presence of at least one of the following ICD9 codes (410, 411, 412, 413, 41, V45.81) or ICD10 codes (I21, I22, I24, Z95.1, Z98.61, or I20.0)^15^.

We calculated age per participant as of August 1, 2022 and determined sex using self-reported sex at birth information provided in AoU. Current smoking status, family history of T2D, self-reported race, and self-reported ethnicity information in AoU were also extracted from participant-provided surveys. We estimated enrichment of sex, family history of T2D, race, ethnicity, and smoking status between T2D cases and non-diabetics using a chi2 test, and tested for significant differences in average values for age and the other T2D-relevant quantitative values using a t-test.

### Genetic Data Curation

#### All of Us

Quality control steps for AoU srWGS data has previously been described in detail^35^. Briefly, AoU removed low quality variants based on a GQ threshold of <20, a DP threshold of <10, an allele balance filter of <0.2, a low QUAL score (60 for SNPs and 69 for indels), or an ExcessHet score <54.69. Rare variants, defined as having a minor allele count <100 or minor allele frequency <1% in all genetically-inferred populations, were also removed. AoU also provides a list of individuals flagged as having low quality WGS data based on the number of variants, insertion/deletion ratio, number of insertions, number of deletions, variant and indel heterozygosity, transition/transversion ratio, and the number of variants not in gnomAD v3.1, which we removed from our analysis. AoU also previously flagged a minimum set of individuals with kinship >0.1 to another individual, which we also used to remove related participants.

Genetically-inferred population groupings of participants were previously performed using a random forest classifier and reference populations from 1000 Genomes (1000G), the Human Genome Diversity Project, and gnomAD^35^. Population labels were then defined using a 75% probability cutoff. Following guidelines for reporting Race, Ethnicity, and Ancestry^36^, we here refer to individuals in AoU who are assigned as being genetically similar to the reference populations from Europe, Africa, and the Americas as EUR-like, AFR-like, and AMR-like respectively^22,23^. To obtain genetic principal components (PCs) in AoU, we first ran PCA on all participants in 1000G, then projected AoU participants into the same PC space.

#### Penn Medicine Biobank

Detailed information on genotyping and imputation for PMBB can be found in a previous publication^37^. Briefly, participants were genotyped using a custom genotyping array constructed from the Global Screening Array or genotype by sequencing capture. SNPs with more than 5% missingness were excluded, and samples with over 10% missingness or sex discrepancies were also removed using PLINK 2.0. Following genotype quality control, imputation was conducted using the TOPMed version R3 reference panel via the Michigan TOPMed Imputation Server 2 with minimac v4.1.6. Phasing was carried out using Eagle v2.4 and the GRCh38/hg38 genome build. Variants were retained if they were in one of the genotype arrays or had an average R2 greater than 0.2.

PMBB previously inferred participants’ genetic similarity to reference populations. To do so, they performed PCA on unrelated individuals based on kinship up to the 3^rd^ degree with the *smartpca* module from the EIGENSOFT package on an 182,701 common SNPs previously pruned for LD, MAF <0.1, and hardy-weinberg equilibrium P<1e-6. They then clustered the study population based on genetic similarity to a superpopulation from 1000G using quantitative discriminant analysis (QDA) based on the top 20 principal components. We only included EUR-like individuals in the analysis.

### PGS Estimation

We estimated novel single-population PGS for T2D, BMI, and CHD using PRS-CS with default parameters^19^ and 1000G reference panels provided in the PRS-CS GitHub. We also used previously published GWAS for T2D and BMI performed in single, EUR-like population groups^24,31^, as well as a mostly EUR-like recent meta-analysis for CHD^15^. We chose to focus on EUR-like populations initially since they were the majority single population in AoU and had the highest sample sizes in available GWAS. We also created a multi-ancestry PGS for T2D using PRS-CSx with default parameters^28^ and the --meta flag, using GWAS from five different populations (AFR-like, EUR-like, AMR-like, EAS-like, and SAS-like)^24^.

PRS-CS and PRS-CSx both use a Bayesian approach to estimate posterior effects per variant to be included in the PGS based on input GWAS summary statistics and LD reference panels via and MCMC step, averaging the posterior samples per MCMC step to calculate the final posterior effect. However, previous work has demonstrated that the posterior samples per MCMC step can also be used to estimate the full posterior distribution of a variant’s effect size, and thus a posterior distribution on an individual’s PGS^13^. This posterior distribution can then be used to capture uncertainty in PGS estimation per individual. To estimate this individual-level PGS, we first extracted 100 posterior effects samples per variant from the MCMC iterations using the --write_pst argument in PRS-CS and PRS-CSx. We then applied these 100 different variant estimates into AoU to generate 100 different PGS per individual. Finally, each of the PGS were adjusted for global ancestry and standardized as described above. We used these 100 different PGS to approximate the PGS posterior distribution per individual. As we predict the uncertainty in the PGS from the posterior samples of PRS-CS and PRS-CSx, we refer to these ranges of scores as the “predicted range”. For association testing, we also estimated a single point estimate per individual based on these 100 PGS posterior samples by estimating their individual posterior mean, or PGS point estimate.

We also downloaded 62 PGS for T2D, 96 PGS for BMI, and 48 PGS for CHD from the PGS Catalog^38^, a public collection of published PGS for a variety of traits and diseases. We removed PGS with missing information on non-effect alleles and PGS that were trained in AoU to avoid overfitting, resulting in 57, 83, and 40 PGS for T2D, BMI, and CHD respectively. We then applied these PGS into AoU using PLINK. We also adjusted each of these published PGS for global ancestry and standardized as described above.

As the performance of different published PGS at a population-level varies^8,15^, and to properly compare our new PGS with published PGS of similar accuracy, we further filtered the published scores based on either their area under the receiver operator curve (AUC) for prevalent T2D and CHD or their R2 for BMI. We estimated AUC and R2 values for each published PGS and for the posterior mean of the new PRS-CS and PRS-CSx PGS using logistic or linear regression models, adjusted for age, sex, the first 10 PCs, and sequencing location. We then identified published PGS with at least 95% probability of being equivalent at a population-level using a Region of Practical Equivalence (ROPE) estimate. For our primary analyses, we used a pre-specified range of +/- 0.02, though we also tested +/- 0.04 and +/- 0.01 in sensitivity analyses. To implement the ROPE approach, we used a 10-fold cross validation approach with six repeats with the vfold_cv function from rsample R package. We then performed resampling of the results with the perf_mod function, then selected population-equivalent scores with the contrast_models function from the tidyposterior R package. As the PGS which pass this ROPE criteria are already published, we refer to the range of these published scores as the “empirical range”. We estimated rank concordance between scores using Spearman correlations.

### Relationship between empirical and predicted PGS credible ranges

We define the empirical range of a PGS as the length of the k-th credible range across published PGS. Similarly, we define the predicted range of a PGS as the length of the k-th credible range across posterior samples of a Bayesian-based PGS. As examples, we focus on the 25%, 50%, 75%, 90%, and 95% credible ranges for our analyses.

As a basic simulation, we compared how the empirical and predicted PGS ranges would differ if the underlying variance per range differed as well. To do this, we sampled true, standardized PGS for 10,000 individuals from a gaussian distribution. Since an individual’s estimated PGS is expected to be similar to but not equal to their true PGS, as the true causal effects of each variant are unknown, per individual we re-sampled their estimated PGS from a normal distribution centered at their true PGS and with some pre-specified variance. For the empirical PGS, we sampled 10 PGS per individual with some pre-specified “empirical PGS variance”. This small number of samples reflects the similar number of published, population-equivalent PGS for T2D. Similarly, we simulated predicted PGS credible ranges with 100 sampled PGS per individual with some pre-specified “predicted PGS variance”. Since in practice the number of published PGS is much smaller than the number of posterior samples of the PGS, accurate estimation of the empirical credible range at an individual level is impossible. Instead, we grouped participants into percentile bins based on their Bayesian-based PGS, then estimated the credible range width per bin.

For real data, we repeated the above analysis using the population-equivalent PGS to construct the empirical ranges and the PGS posterior samples from PRS-CS or PRS-CSx to construct the predicted ranges.

### Estimation of predicted credible ranges with PredInterval

PredInterval leverages the CV+ framework to estimate the prediction intervals of a phenotype based on a PGS^27^. To compare the prediction intervals from PredInterval with the credible ranges from the posterior samples of PRS-CS, we first used PUMAS^39^ to re-sample the EUR-like GWAS originally used as input into PRS-CS into three different GWAS, for 3-fold CV. We chose a 3-fold CV to reduce the computational burden of running PredInterval and due to similar prediction interval widths across fold number^27^. Next, we estimated the posterior mean point estimates of the variant weights for each of these three different GWAS independently using PRS-CS, as described above. Since PredInterval requires a calibration dataset, we used newly released EUR-like individuals in the AoU v8 data release but not the AoU v7 data release for calibration, then applied PredInterval to the EUR-like individuals in AoU v7. Similar to above, we also estimated the prediction intervals per percentile bin, as well as per individual.

### Risk agreement and confidence estimation

Total risk agreement across the published, empirical scores per individual was calculated as the average number of published PGS which call said individual as high risk based on the same threshold *t*. We termed this metric as “stability of PGS risk”.

We estimated the probability of an individual being at high risk for a PGS given the uncertainty in their novel PGS estimate as the total proportion of their posterior PGS distribution being over the same risk threshold *t*. We term this probability of being at risk as an individual’s “risk confidence”. Unlike the previous metric of PGS risk “stability” which is estimated from multiple published PGS, this risk confidence metric is from a single, novel PGS not included in the calculation of “stability”. As such, it represents the probability of being high risk based on only the novel PGS when accounting for potential variations in how the variants in this PGS are weighted. For further comparisons, we also binned individuals based on their risk confidence values into high confidence (risk confidence >95%), medium-high confidence (95%>= risk confidence >75%), and medium confidence (75% >= risk confidence > 50%). We tested for significant differences in average stability across confidence groups using pairwise Mann-Whitney U tests.

### Longitudinal Analyses

Before running time-to-event analyses, individuals were filtered to include those with at least 3 outpatient visits with a primary care physician. We then constructed a longitudinal cohort from the EHR data as described previously^11^. Participants were included if they their first outpatient visit between January 1, 2000 and April 1, 2023. As data collection for AoU ended in April 2023, this allowed for a maximum of 23 years. Time-to-event was then calculated as the difference in time between the first outpatient visit and either the first T2D ICD-code or April 2023. We further censored participants if they had a T2D ICD code prior to their first recorded outpatient visit or within six months after their first recorded outpatient visit. In total, this left us with 144,516 participants, including 21,437 and 123,079 patients with and without T2D respectively.

We computed Kaplan-Meier curves and ran cox models using the lifelines Python package. For cox models, we pulled known clinical risk factors for T2D, including age, sex, smoking status, family history of T2D, BMI, systolic blood pressure, total cholesterol, HDL cholesterol, triglycerides, and random glucose. We calculated age as of the patient’s first outpatient visit, and cleaned quantitative values to remove outliers more than three standard deviations away from the average in log space. We then only considered quantitative values measured up to 1.5 years prior to the patient’s first outpatient visit and 0.5 years after the patient’s first outpatient visit, taking the average if multiple measurements were available. Due to the low number of repeated BMI measurements, we used the average BMI without time restraints in our analyses as described previously^11^. We estimated enrichment of sex, family history of T2D, race, ethnicity, and smoking status between confidence groups using a chi2 test, and tested for significant differences in average values for age and the other T2D-relevant quantitative values using a One-way Anova test.

In addition to these clinical risk factors, we also included the first 10 PCs as covariates in the cox models. Cox models also included either the posterior mean value of the PGS alone, or both the posterior mean value of the PGS and high risk confidence information, to evaluate the additional improvements in model prediction due to high risk confidence. In total, this left us with 7,233 participants with all clinical risk factors measured, including 852 with T2D and 6,381 without T2D. Absolute probabilities for developing T2D were estimated using logistic regression models fit on these 7,233 participants, including the same covariates as the cox models above. The posterior mean value of the PGS and risk confidence were also included in the models as quantitative, standardized values.

## Data Availability

Individual-level phenotype and genetic data from All of Us are available on the All of Us Research Workbench for all approved, controlled-tier researchers. Applied phenotyping algorithms and PGS within All of Us can be made available to other approved All of Us researchers upon request. Individual-level data from Penn Medicine BioBank cannot be shared to researchers without an approved Penn Medicine BioBank IRB, but summary statistics can be made available upon reasonable request. Novel PGS will be made publicly available upon publication.

## Acknowledgements

We gratefully acknowledge *All of Us* participants for their contributions, without whom this research would not have been possible. We also thank the National Institutes of Health’s All of Us Research Program for making available the participant data examined in this study.

We also acknowledge the Penn Medicine BioBank (PMBB) for providing data and thank the patient-participants of Penn Medicine who consented to participate in this research program. We would also like to thank the Penn Medicine BioBank team and Regeneron Genetics Center for providing genetic variant data for analysis. The PMBB is approved under IRB protocol #813913 and supported by Perelman School of Medicine at University of Pennsylvania, a gift from the Smilow family, and the National Center for Advancing Translational Sciences of the National Institutes of Health under CTSA award number UL1TR001878.

## Author Contributions

R.M. performed the primary analyses for T2D in AoU, compared stability and confidence for BMI and CHD, designed experiments and analyses, and created the figures. Z.S. downloaded and applied CHD PGS from the PGScatalog, and Z.S. and S.A.A. generated novel CHD PGS with uncertainty information. X.L. downloaded and applied BMI PGS from the PGScatalog, and generated novel BMI PGS with uncertainty information. S.L. performed T2D phenotyping within PMBB. R.M. applied the published and novel T2D PGS into PMBB, and compared credible range sizes between the published and predicted PGS. M.G.L and S.M.D. provided feedback throughout the project and revised the paper. R.M. and B.P. conceived and planned the study, supervised, and coordinated the analyses, and wrote the manuscript.

