## Supplementary Material for "Statistical uncertainty explains the poor agreement in polygenic scoring for type 2 diabetes"

**Table of Contents**


| | |
|---|---|
| **Penn Medicine Biobank Team and Contributions** | **2** |
| **Supplementary Figure 1** | **4** |
| **Supplementary Figure 2** | **5** |
| **Supplementary Figure 3** | **6** |
| **Supplementary Figure 4** | **7** |
| **Supplementary Figure 5** | **8** |
| **Supplementary Figure 6** | **9** |
| **Supplementary Figure 7** | **10** |
| **Supplementary Figure 8** | **11** |
| **Supplementary Figure 9** | **11** |
| **Supplementary Figure 10** | **12** |
| **Supplementary Figure 11** | **13** |
| **Supplementary Figure 12** | **14** |
| **Supplementary Figure 13** | **16** |
| **Supplementary Figure 14** | **18** |
| **Supplementary Figure 15** | **19** |
| **Supplementary Figure 16** | **19** |
| **Supplementary Figure 17** | **20** |

***Penn Medicine BioBank Team and Contributions***

**PMBB Leadership Team**

Daniel J. Rader, M.D., Marylyn D. Ritchie, Ph.D.

**Contribution**: All authors contributed to securing funding, study design and oversight. All authors reviewed the final version of the manuscript.

**Patient Recruitment and Regulatory Oversight**

JoEllen Weaver, Nawar Naseer, Ph.D., M.P.H., Giorgio Sirugo, M.D., P.h.D., Afiya Poindexter, Yi-An Ko, Ph.D., Kyle P. Nerz

**Contributions**: JW manages patient recruitment and regulatory oversight of study. NN manages participant engagement, assists with regulatory oversight, and researcher access. GS assists with researcher access. AP, YK, KPN perform recruitment and enrollment of study participants.

**Lab Operations**

JoEllen Weaver, Meghan Livingstone, Fred Vadivieso, Stephanie DerOhannessian, Teo Tran, Julia Stephanowski, Salma Santos, Ned Haubein, P.h.D., Joseph Dunn

**Contribution**: JW, ML, FV, SD conduct oversight of lab operations. ML, FV, AK, SD, TT, JS, SS perform sample processing. NH, JD are responsible for sample tracking and the laboratory information management system.

**Clinical Informatics**

Anurag Verma, Ph.D., Colleen Morse Kripke, M.S. DPT, MSA, Marjorie Risman, M.S., Renae Judy, B.S., Colin Wollack, M.S.

**Contribution**: All authors contributed to the development and validation of clinical phenotypes used to identify study subjects and (when applicable) controls.

**Genome Informatics**

Anurag Verma Ph.D., Shefali S. Verma, Ph.D., Scott Damrauer, M.D., Yuki Bradford, M.S., Scott Dudek, M.S., Theodore Drivas, M.D., Ph.D.,

**Contribution**: AV, SSV, and SD are responsible for the analysis, design, and infrastructure needed to quality control genotype and exome data. YB performs the analysis. TD and AV provides variant and gene annotations and their functional interpretation of variants.

For PMBB, please use:

For Regeneron, please use:

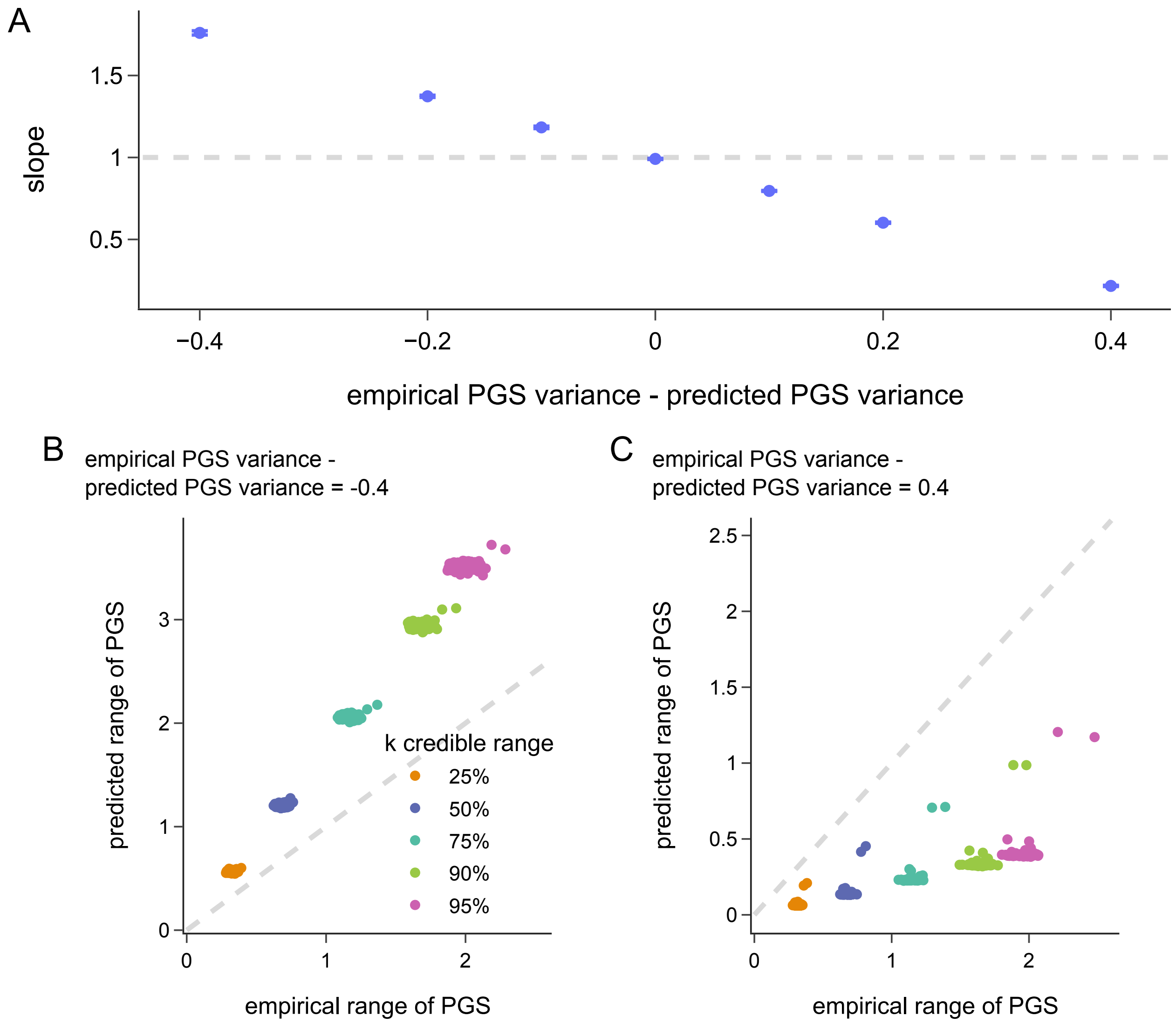


***Supplementary Figure 1: Simulation of differences between predicted and empirical PGS ranges.*** *(**A**) Estimated regression slopes between empirical range and predicted range of a PGS under different conditions where variance in the predicted range is greater than, equal to, or less than the variance in the empirical range. Per simulation, 10,000 individuals were simulated with a true genetic risk from a standard normal distribution, and empirical and predicted PGS per individual were independently sampled from two normal distributions centered at the true genetic risk with variance corresponding to the empirical or predicted range. Individuals were further binned based on percentile of their true genetic risk for accurate*

*credible range calculation (**Methods**). (**B**)-(**C**) Examples of single simulation results when the predicted range variance is less than (**B**) or more than (**C**) the empirical range variance.*

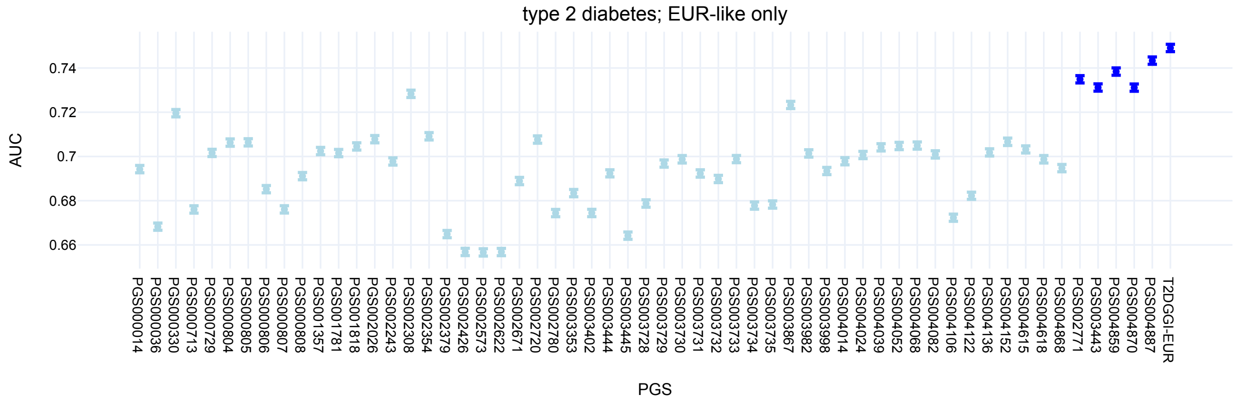


***Supplementary Figure 2: Population-level performance of T2D PGS in AoU.*** *Area under the receiver operator curves for different PGS for T2D within individuals genetically similar to European populations in All of Us. Dark blue points represent PGS with similar AUC based on a region of practical equivalence of 0.02 to the T2DGGI-EUR PGS. Error bars represent the 95% confidence interval based on bootstrapping.*

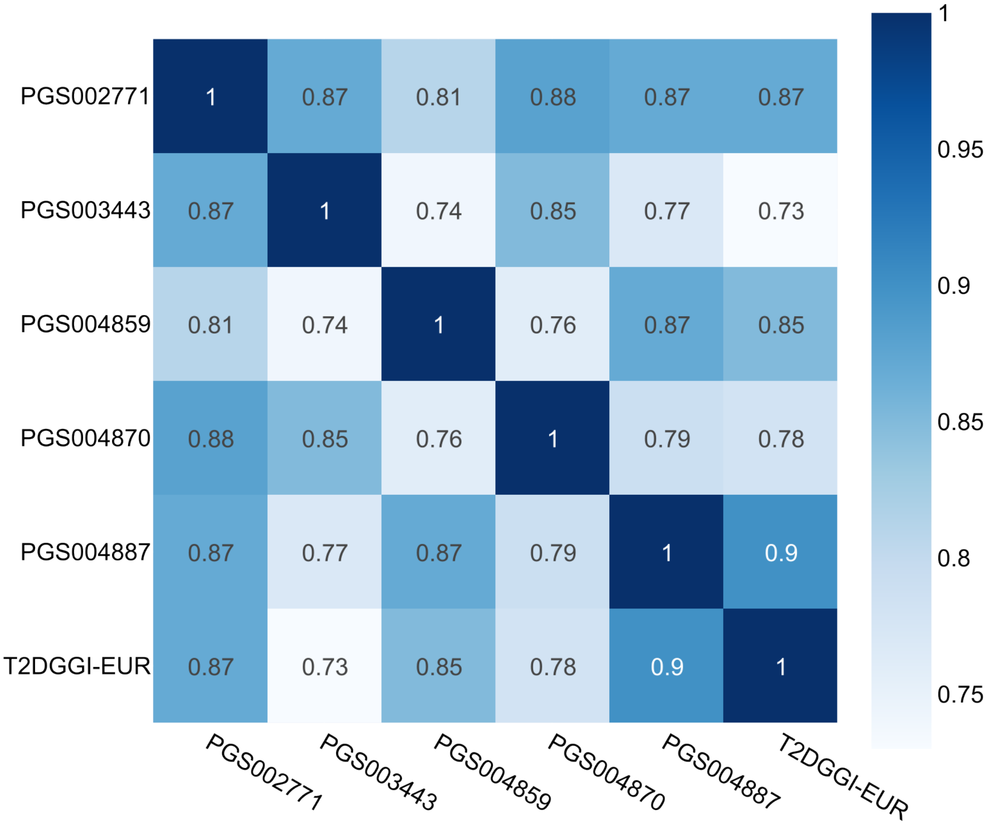


***Supplementary Figure 3: Rank concordance of population-level equivalent T2D PGS.***

*Heatmap of pairwise Spearman correlation coefficients between different T2D PGS with area under the receiver operator curve overlapping within a region of practical equivalence margin of 0.02 to the T2DGGI-EUR score, considering only individuals genetically similar to European populations in All of Us.*

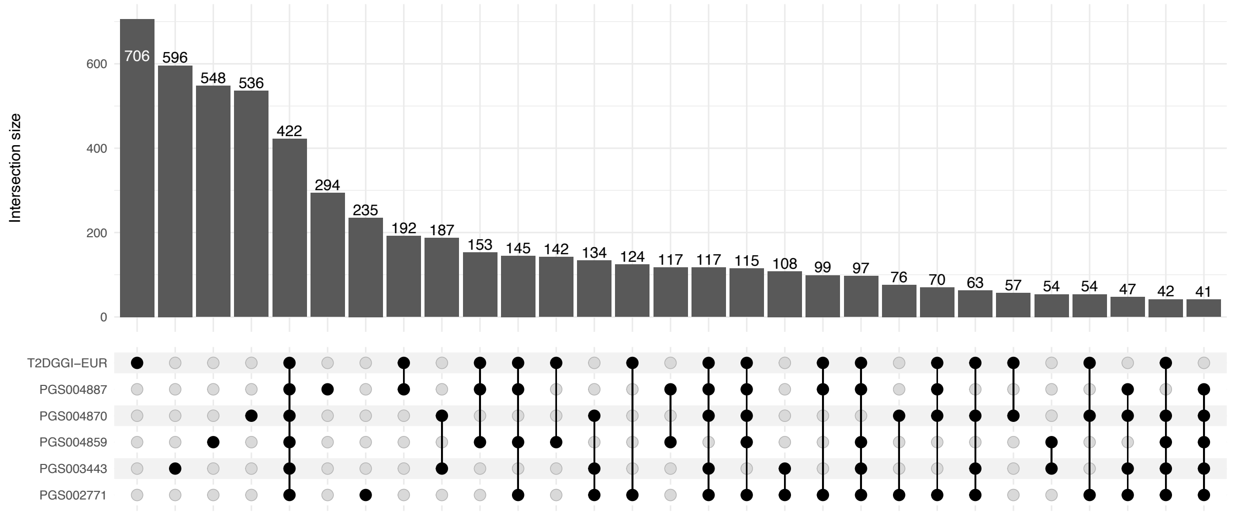


***Supplementary Figure 4: Agreement of T2D PGS risk classification with a threshold of top 2%.*** *Upset plot of the overlapping number of individuals identified as high risk between different published T2D PGS, with area under the receiver operator curve overlapping within a region of practical equivalence margin of 0.02 to the T2DGGI-EUR score in All of Us. We defined high risk using the recommended top 2% threshold from the eMERGE consortium. The plot only includes individuals genetically similar to European populations.*

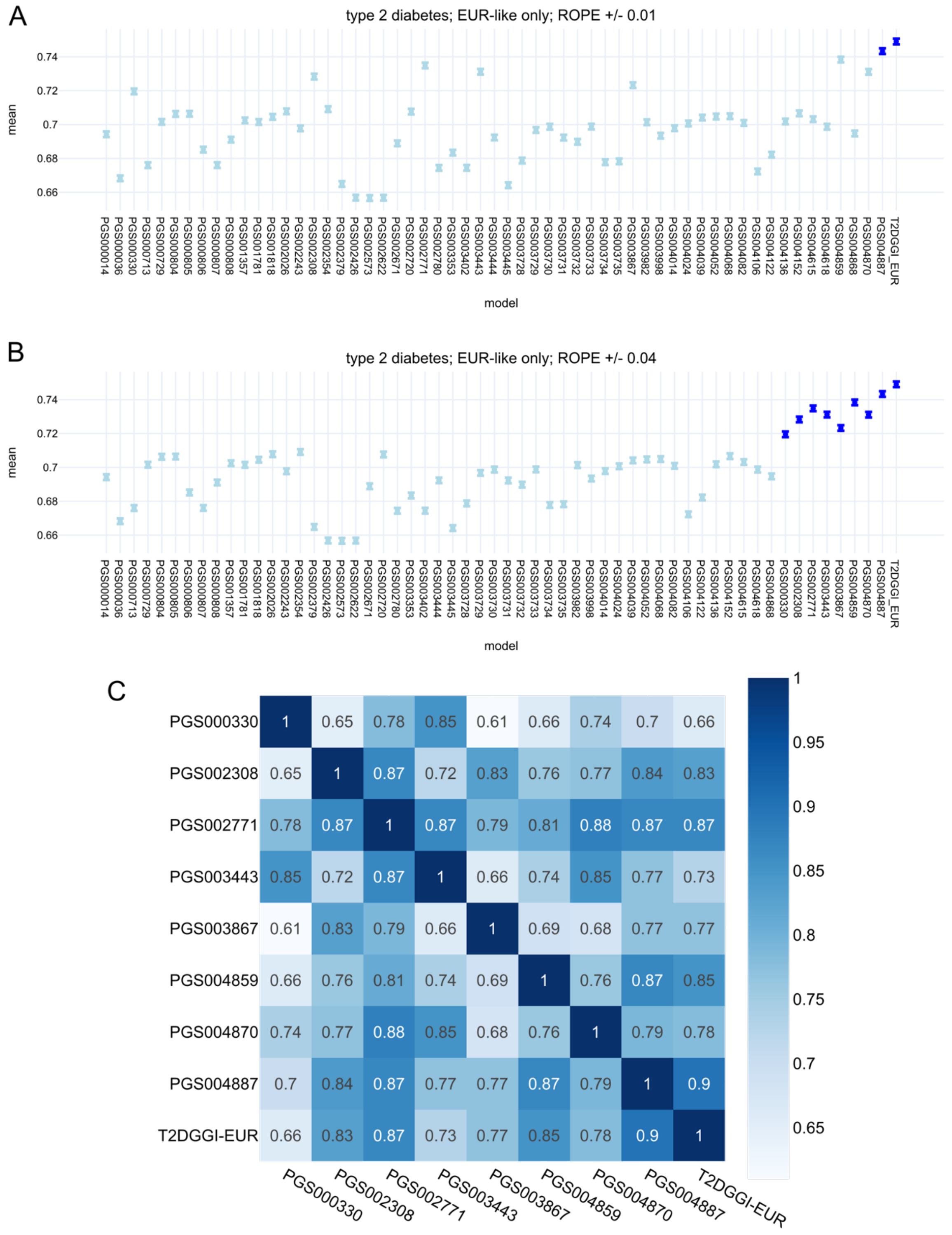


***Supplementary Figure 5: Agreement of T2D PGS with different ROPE cutoffs.*** *(**A**)-(**B**) Area under the receiver operator curves (AUC) for different PGS for T2D within individuals genetically*

*similar to European populations in All of Us. Dark blue points represent PGS with similar AUC based on either a region of practical equivalence (ROPE) +/- 0.01 (**A**) or +/- 0.04 (**B**) to the T2DGGI-EUR. Error bars represent the 95% confidence interval based on bootstrapping. (**C**) Heatmap of pairwise Spearman correlation coefficients between different T2D PGS with population-equivalence to the T2DGGI-EUR PGS based on a ROPE +/- 0.04.*

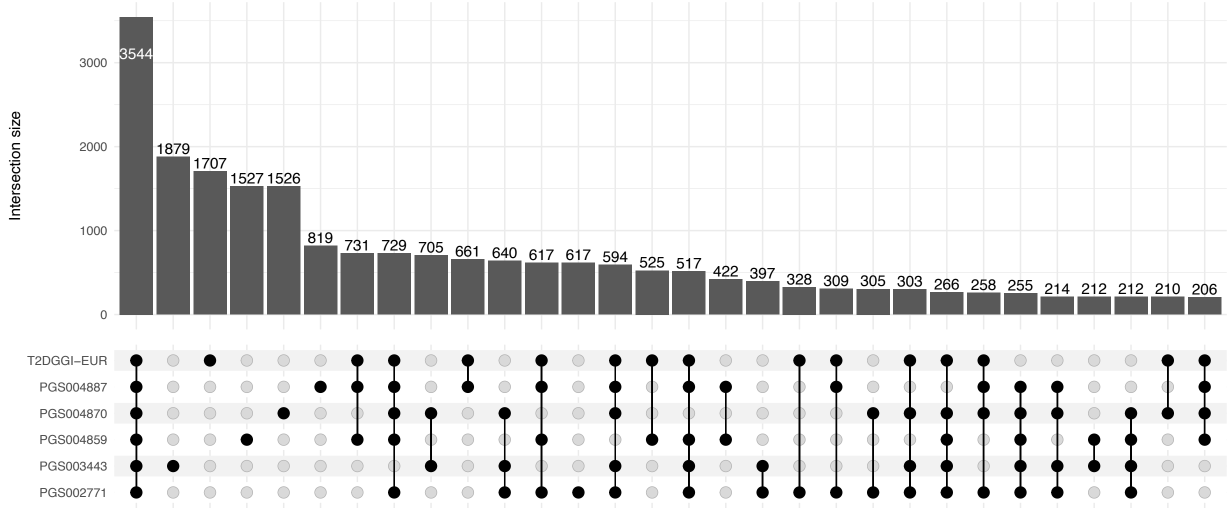


***Supplementary Figure 6: Agreement of T2D PGS risk classification with a threshold of top 10%.*** *Upset plot of the overlapping number of individuals identified as high risk between different type 2 diabetes PGS with area under the receiver operator curve overlapping within a region of practical equivalence (ROPE) +/- 0.02 to the T2DGGI-EUR score in All of Us. We defined high risk using a lenient top 10% threshold. The plot only includes individuals genetically similar to European populations.*

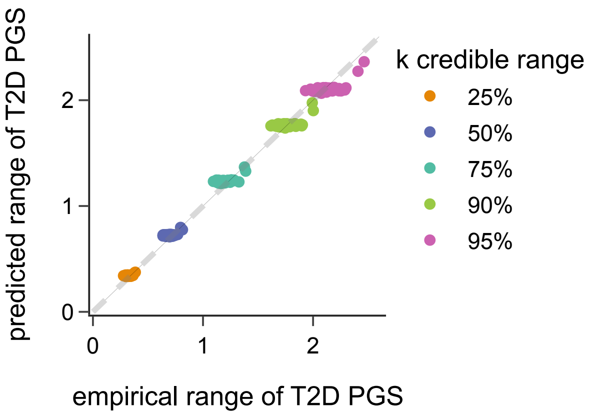


***Supplementary Figure 7: Replication of calibrated predicted T2D PGS ranges.*** *Scatterplot of the empirical range of T2D PGS based on published scores to predicted range based on the T2DGGI-EUR score from PRS-CS, across different credible range sizes in individuals genetically similar to European populations in the Penn Medicine Biobank. We select published scores included in the empirical ranges with a region of practical equivalence (ROPE) +/- 0.02 around the T2DGGI-EUR area under the receiver operator curve based on the results in All of Us (**Supplementary Table 3**). Due to the small number of published T2D PGS used, we grouped individuals into percentile bins prior to range calculation (**Methods**).*

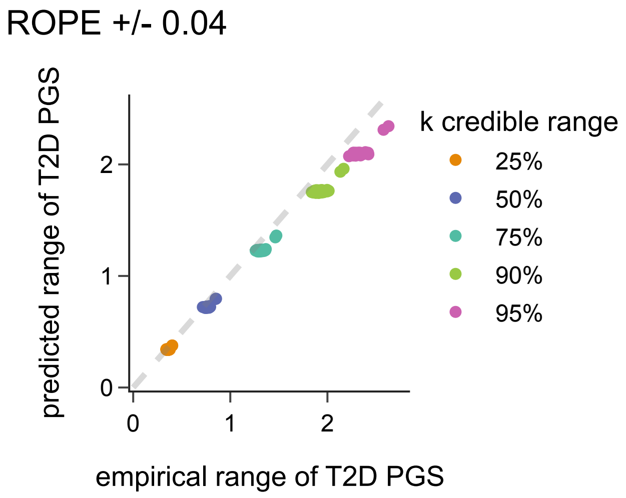


***Supplementary Figure 8: Calibration of predicted and empirical T2D PGS ranges varies with empirical range definition.*** *Scatterplot of the empirical range of T2D PGS based on published scores to predicted range based on the T2DGGI-EUR score from PRS-CS, across different credible range sizes in All of Us. We select published scores included in the empirical ranges with a region of practical equivalence (ROPE) +/- 0.04 around the T2DGGI-EUR area under the receiver operator curve. Due to the small number of published T2D PGS used, we grouped individuals into percentile bins prior to range calculation (**Methods**).*

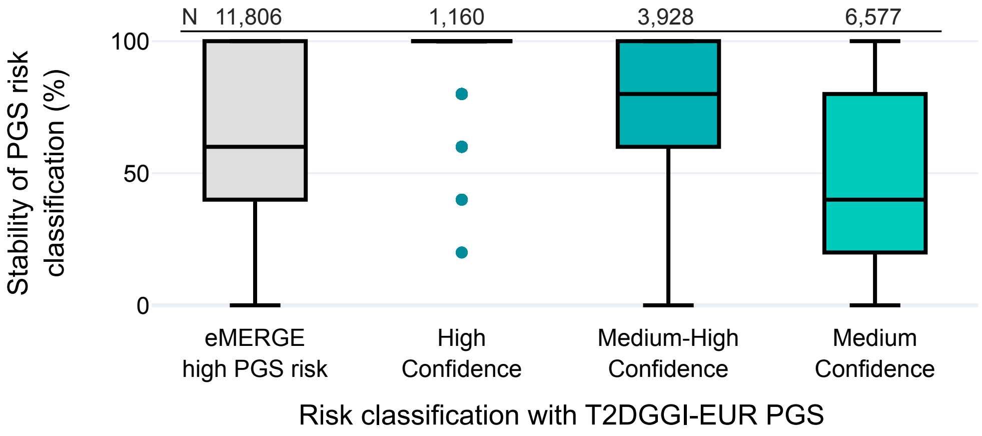

***Supplementary Figure 9: PGS risk agreement associates with risk confidence at a risk cutoff of top 10%.*** *Comparison of risk stability across individuals identified using a risk cutoff of top 10% with the T2DGGI-EUR score and individuals identified at different confidence classifications with the T2DGGI-EUR score in All of Us. Stability per individual was calculated as the average number of PGS that call said individual as high risk.*

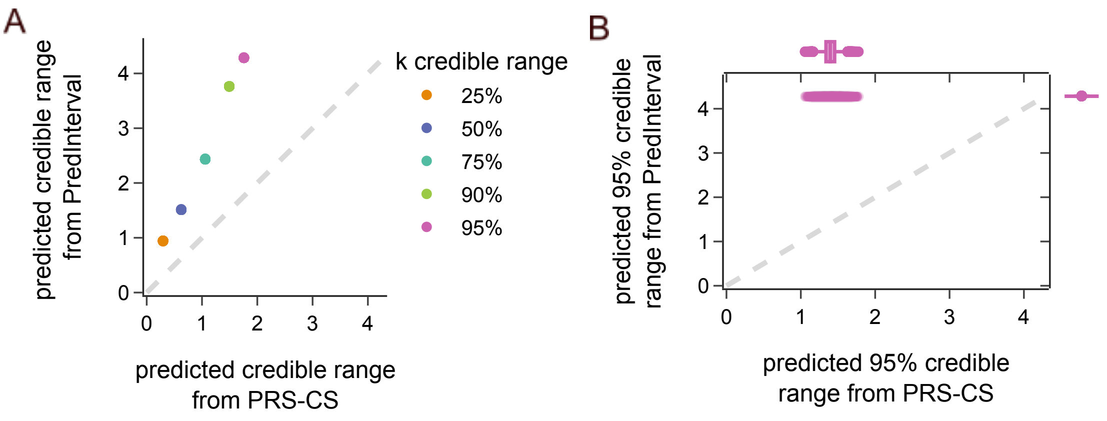


***Supplementary Figure 10: Comparison of credible ranges estimated directly from PRS-CS and PredInterval.*** *(**A**) Scatterplot of the predicted range of the T2D PGS based on the T2DGGI-EUR score posterior distribution from PRS-CS compared to the predicted range from PredInterval, across different credible range sizes in All of Us. For consistency with previous plots, we grouped individuals into percentile bins prior to range calculation (**Methods**). (**B**) Marginal scatter and box plot of the predicted 95% credible range per individual estimated based on the T2DGGI-EUR score posterior distribution from PRS-CS compared to the predicted 95% credible range of the T2DGGI-EUR score from PredInterval.*

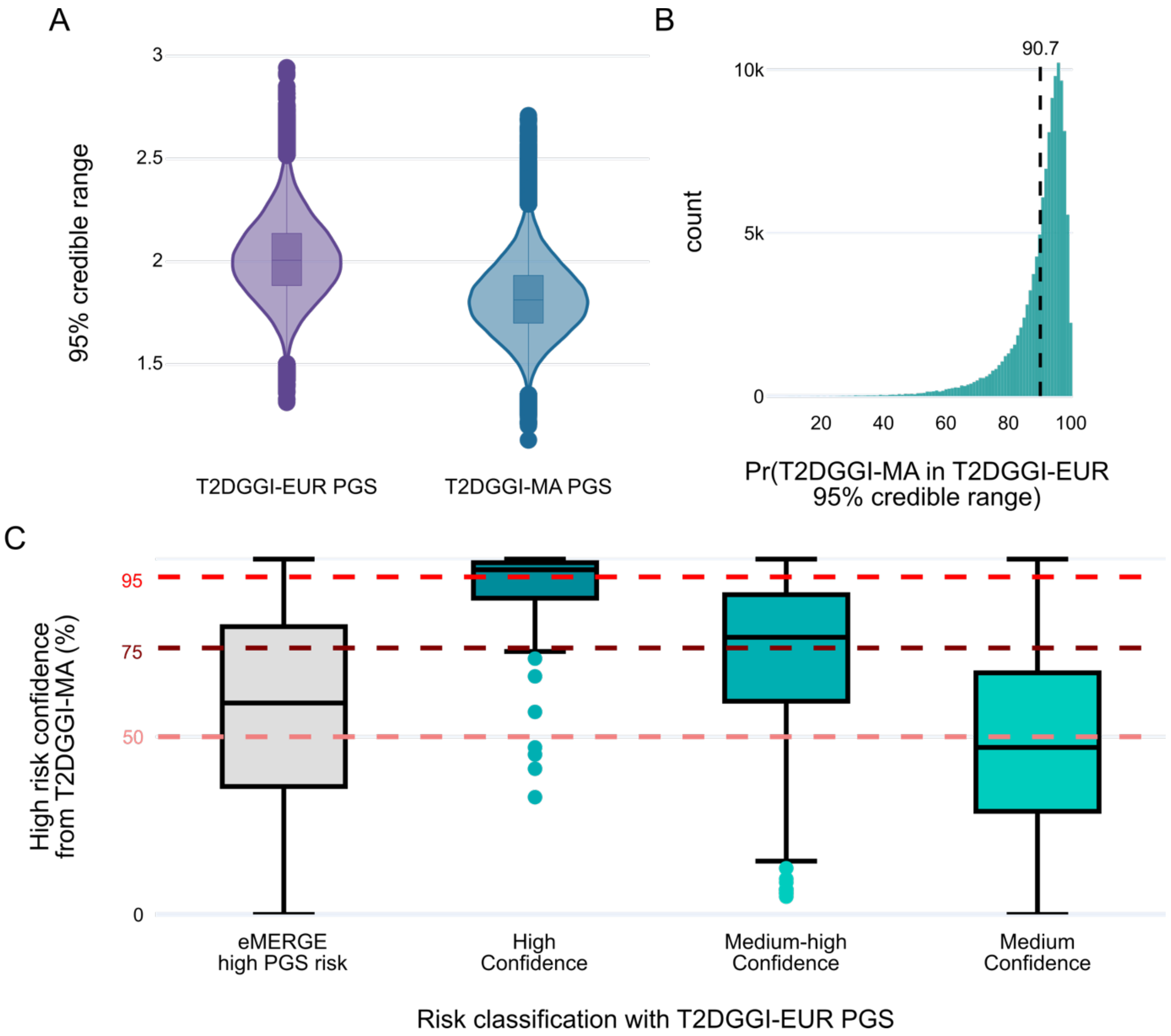


***Supplementary Figure 11: Concordance between risk confidence as estimated from two separate PGS.*** *(**A**) Comparison of the 95% credible range sizes between a multi-ancestry type 2 diabetes PGS (T2DGGI-MA) and a European type 2 diabetes PGS (T2DGGI-EUR). (**B**) Histogram of the probability of an individual's T2DGGI-MA PGS being within their T2DGGI-EUR PGS 95% credible range. The black dashed line represents the mean probability. (**C**) Confidence of being above the high PGS risk threshold of top 2% based on the T2DGGI-MA PGS compared to different risk classifications schemas using the T2DGGI-EUR PGS. Dashed red lines indicate the confidence cutoffs used to define individuals as high confidence (>95%),*

*medium-high confidence (>75%), and medium confidence (>50%). Only individuals genetically similar to European populations in All of Us were included in the analysis.*

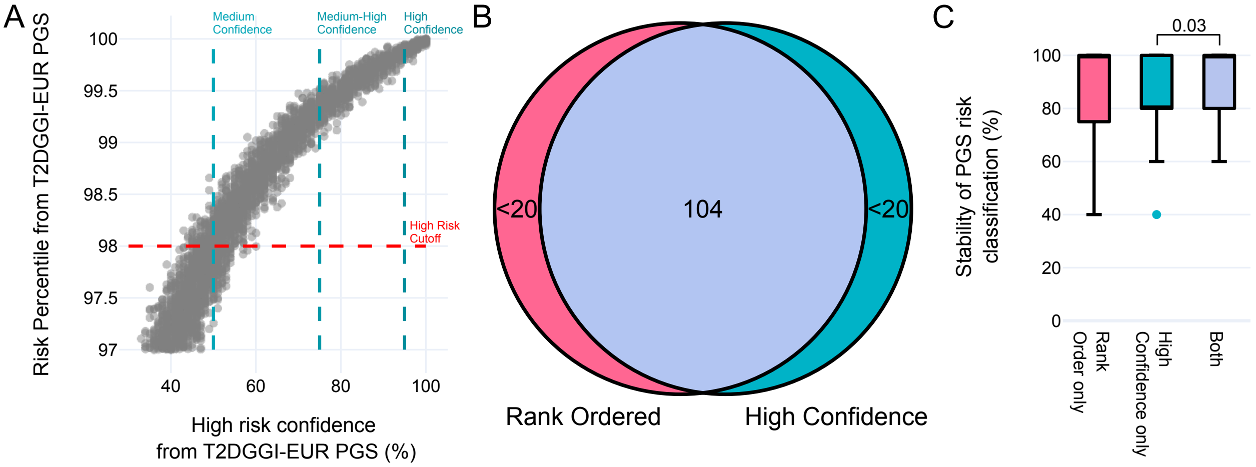


***Supplementary Figure 12: Metrics of high PGS risk confidence correlate with singular point estimates of PGS.*** *(**A**) Scatter plot of confidence of being above the top 2% risk threshold based on the T2DGGI-EUR PGS compared to the single point estimate of risk based on the same PGS. Dashed red horizontal line represents the top 2% risk threshold, and the vertical blue dashed lines represent different high risk confidence cutoffs used to define medium confidence, medium-high confidence, and high confidence groups. (**B**) Overlap of individuals identified as high confidence of risk based on the T2DGGI-EUR PGS and the same number of individuals identified as having the highest single point estimate of risk based on the same PGS. This latter approach for identifying individuals is referred to as "Rank Ordered". Numbers of participants <20 are censored in accordance with data reporting guidelines from AoU. (**C**) Comparison of PGS risk stability across the 5 published type 2 diabetes PGS with population equivalence to the T2DGGI-EUR PGS based on a region of practical equivalence (ROPE) +/- 0.02 for individuals identified only with the rank ordered approach, only with the high confidence approach, or with both. Nominally significant P-values (P<0.05) based on a Mann-Whitney U*

*test are indicated. Only individuals genetically similar to European populations in All of Us were included in the analysis.*

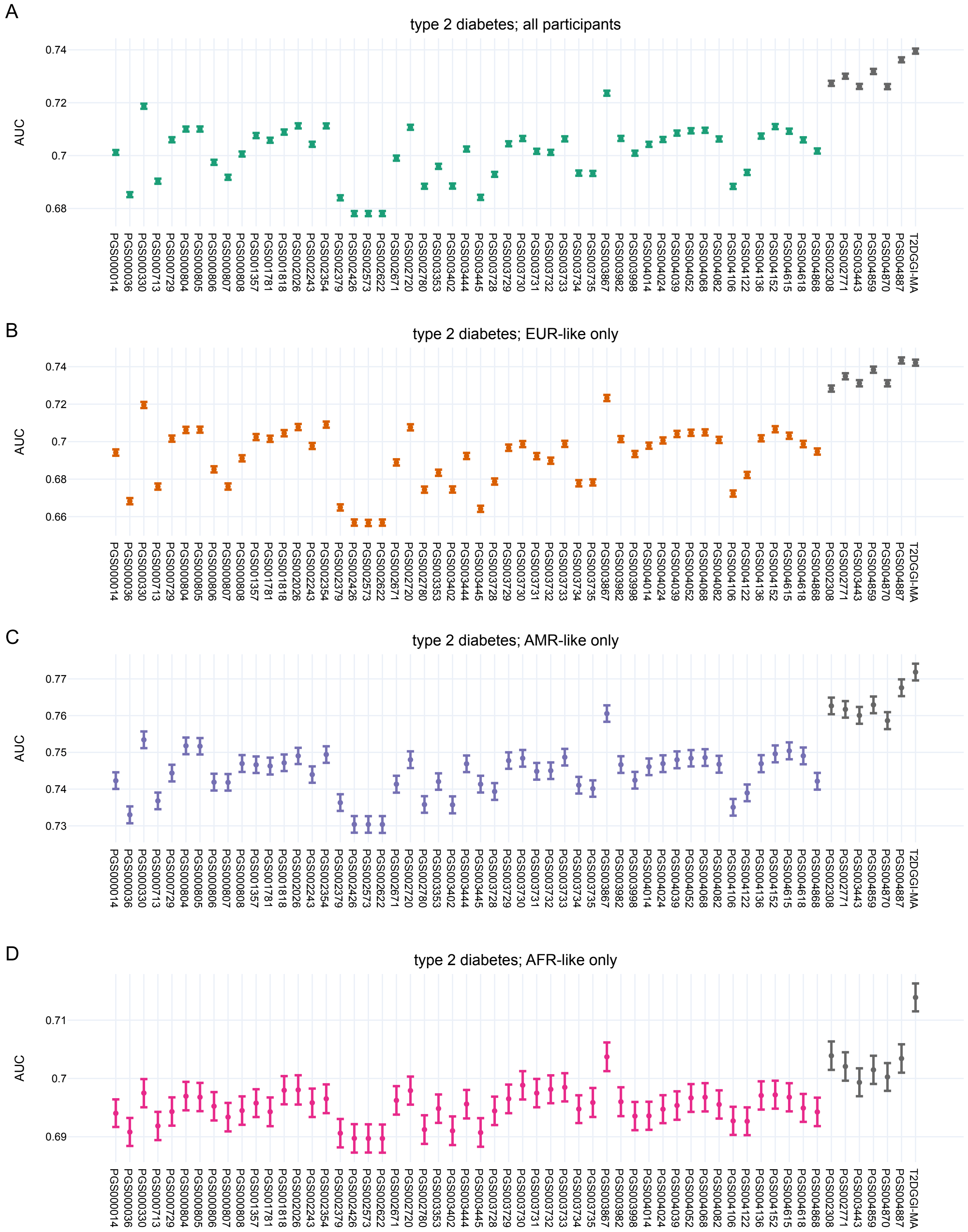
A
type 2 diabetes; all participants
AUC
0.74
0.72
0.7
0.68
PGS000014 PGS000036 PGS000330 PGS000713 PGS000729 PGS000804 PGS000805 PGS000806 PGS000807 PGS000808 PGS001357 PGS001781 PGS001818 PGS002026 PGS002243 PGS002354 PGS002379 PGS002426 PGS002573 PGS002622 PGS002671 PGS002720 PGS002780 PGS003353 PGS003402 PGS003444 PGS003445 PGS003728 PGS003729 PGS003730 PGS003731 PGS003732 PGS003733 PGS003734 PGS003735 PGS003867 PGS003982 PGS003998 PGS004014 PGS004024 PGS004039 PGS004052 PGS004068 PGS004082 PGS004106 PGS004122 PGS004136 PGS004152 PGS004615 PGS004618 PGS004868 PGS002308 PGS002771 PGS003443 PGS004859 PGS004870 PGS004887 T2DGGI-MA
B
type 2 diabetes; EUR-like only
AUC
0.74
0.72
0.7
0.68
0.66
C
type 2 diabetes; AMR-like only
AUC
0.77
0.76
0.75
0.74
0.73
D
type 2 diabetes; AFR-like only
AUC
0.71
0.7
0.69

***Supplementary Figure 13: Population-level performance of type 2 diabetes PGS across different population groups.*** *Area under the receiver operator curves for different PGS for type 2 diabetes within all individuals in All of Us (**A**), individuals genetically similar to European populations (**B**; EUR-like), individuals genetically similar to Admixed American populations (**C**; AMR-like), and individuals genetically similar to African populations (**D**; AFR-like). Grey points represent PGS with similar AUC based on a region of practical equivalence (ROPE) +/- 0.02 to the multi-ancestry T2DGGI-MA PGS in every population group. Error bars represent the standard deviation based on bootstrapping.*

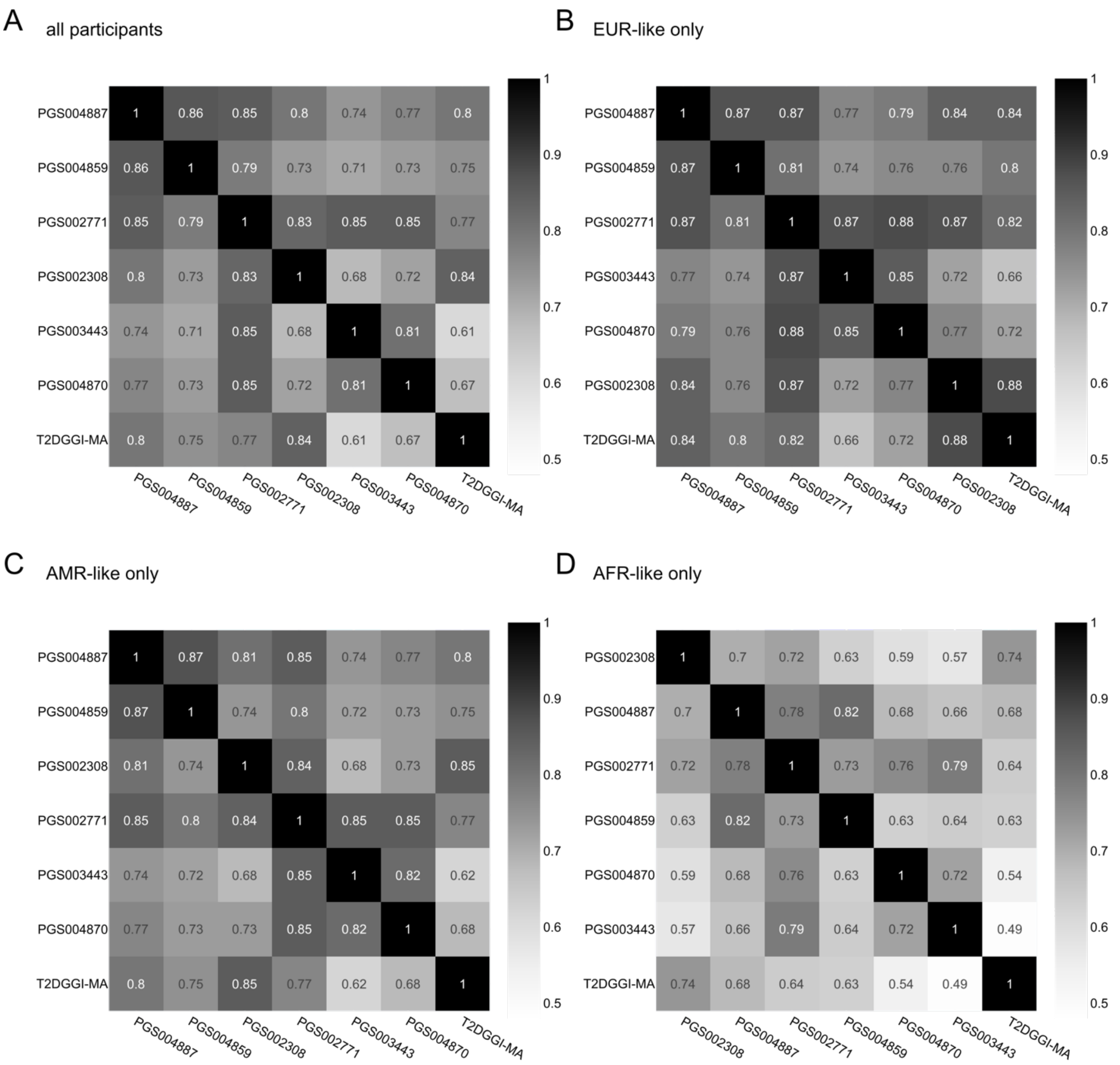


***Supplementary Figure 14: Rank concordance of population-level equivalent type 2 diabetes (T2D) PGS in multiple populations.*** *Heatmaps of pairwise Spearman correlation coefficients between different T2D PGS with area under the receiver operator curve overlapping within a region of practical equivalence margin (ROPE) +/- 0.02 to that of the T2DGGI-MA score in All of Us. Analyses included (**A**) all participants, (**B**) individuals genetically similar to European populations (EUR-like only), (**C**) individuals genetically similar to Admixed American populations (AMR-like only), (**D**) individuals genetically similar to African populations (AFR-like only).*

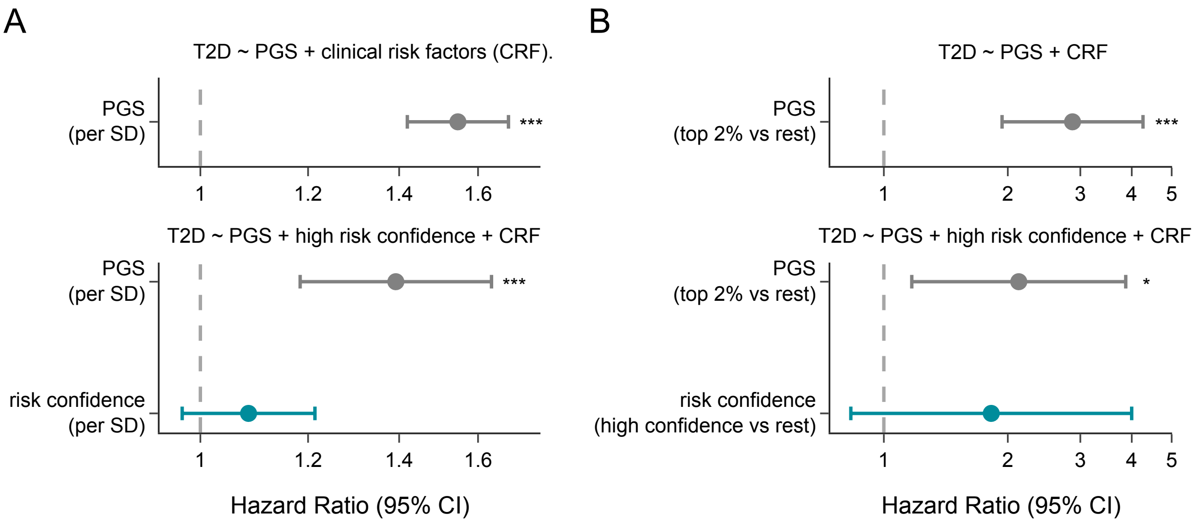


***Supplementary Figure 15: T2D PGS risk confidence is not associated with incident T2D after adjusting for the PGS.*** *(**A**) Hazard ratios of the T2DGGI-MA PGS and risk confidence estimated from the T2DGGI-MA score in joint or PGS-only models in All of Us. Models included previously established T2D clinical risk factors[30] of age, sex, BMI, smoking status, family history of T2D, systolic blood pressure, random glucose, total cholesterol, high-density lipoprotein, and triglycerides. Both the PGS and risk confidence information were quantative and standardized to units per standard deviation (SD). (**B**) Hazard ratios similar to (**A**), however with binary encodings of the PGS and risk confidence information based on the eMERGE risk guidelines (top 2%) and our previous definition of high risk confidence (>95%) respectively. * P<0.05, *** P<0.001.*

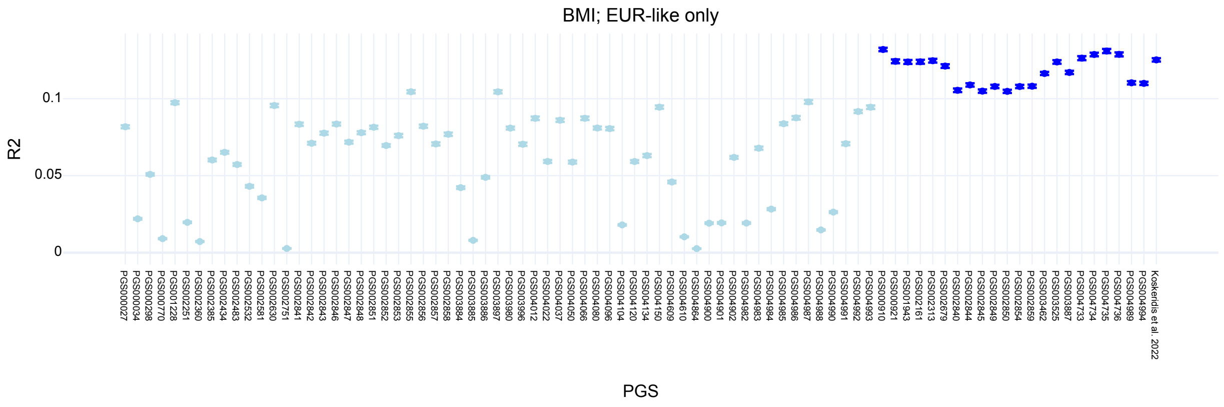

***Supplementary Figure 16: Population-level performance of BMI PGS in AoU.*** *R2 for different PGS for BMI within individuals genetically similar to European populations in All of Us. Dark blue points represent PGS with similar R2 based on a region of practical equivalence of 0.02 to the Koskeridis et al. 2022 score. Error bars represent the standard deviation based on bootstrapping.*

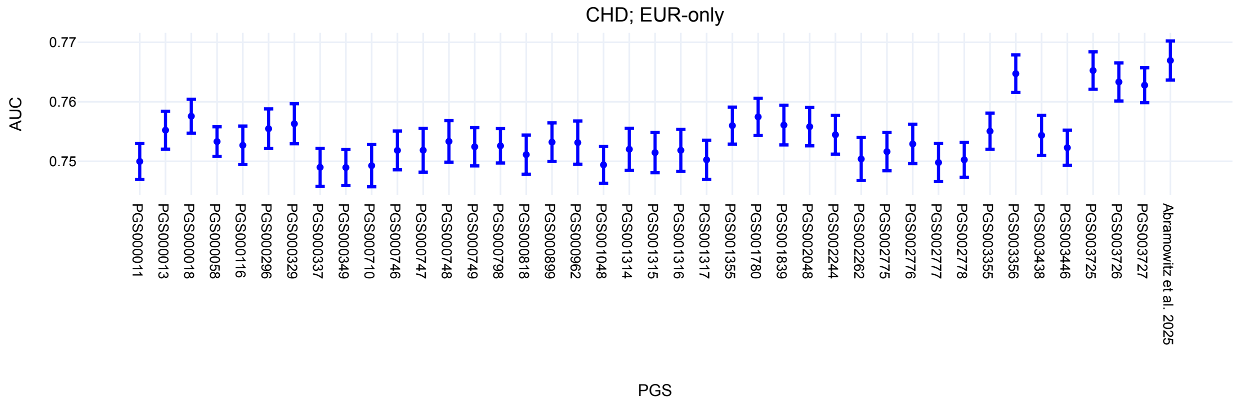


***Supplementary Figure 17: Population-level performance of CHD PGS in AoU.*** *Area under the receiver operator curves for different PGS for CHD within individuals genetically similar to European populations in All of Us. Dark blue points represent PGS with similar AUC based on a region of practical equivalence of 0.02 to the Abramowitz et al. 2025 PGS, which is all tested scores. Error bars represent the standard deviation based on bootstrapping.*
